# Material usage in commercial lateral flow assay kits

**DOI:** 10.1101/2024.07.12.24310277

**Authors:** Marie-Louise Wöhrle, Alice Street, Maïwenn Kersaudy-Kerhoas

**Affiliations:** University of Edinburgh, Edinburgh, EH8 9YL, UK; Heriot-Watt University, Edinburgh EH14 4AS, UK

## Abstract

**Objective:** To assess material usage in commercial lateral flow assay kits and determine baselines for quantitative material requirements in target product profiles for new tests.

**Methods:** We collected and weighted the components of 21 different commercial COVID-19 lateral flow test kits, with emergency use authorisations from the UK MHRA, EU, the US FDA, and the WHO EUL. We took test kits apart manually, classified components and weighted them individually.

**Findings:** Large variations in the total average weights of COVID-19 LFA cassette test kits were observed from 13 g per test to 84 g. The average weight of standard LFA housing in the sample is 4.1 g per casing (range 2.47 g - 6.54 g) whilst the outer packaging makes up between 25% and over 88% of the whole kit and was found to be a large source of weight variations, followed by instructions and LFA housing types.

**Conclusion:** The contribution of LFA tests to global plastic pollution is set to grow year-on-year due to increasingly decentralised testing. Wide variation in the weight of components included in existing COVID-19 test kits suggests there is scope for manufacturers to reduce the amount of materials, including plastic, in test products. We propose that a quantitative baseline of material usage is introduced in future target product profiles for LFA format test kits. This would limit the number of products with a large volume of plastic from reaching the market and reduce the burden of plastic waste from diagnostic testing on local waste management systems.

## 1 Introduction

Rapid testing has become a central pillar of global health, aiding responses to emerging disease outbreaks^1^, expanding universal health coverage^2^, tackling antimicrobial resistance^3^ and contributing to the elimination of neglected diseases^4^. In its 2023 end-of-year report, the Global Fund reported it invested in 53.1 million HIV tests, and 321 million malaria tests. Other sources have reported that the annual production of lateral flow assay (LFA) tests exceeds 2 billion annually^5^. A growing percentage of tests take the format of lateral flow assay devices, and are designed to be used in primary care level (level 1) settings or at home and dispose with the requirement for expensive laboratory infrastructure and expertise. The Global Fund highlights the example of successful investment in increasing HIV at-home tests in Cameroon, Mozambique, Nigeria, Tanzania and Uganda^6^, which brought “*unprecedented development … in the fields of lateral flow technologies*”^7^ (van der Pol 2024, p.135). The associated increased production and spread of single-use plastic cassettes, is leading to considerable waste production and puts environmental health at risk across the countries they are deployed in^8^.

Global health organisations increasingly use Target Product Profiles (TPPs) as a tool to establish consensus among stakeholders around priorities for test design and communicate those needs and expectations to industry^9,10^. By laying out both minimum and optimal requirements for test specifications, TPPs balance existing technical capacities with future aspirations to stimulate innovation. They therefore offer an important point of intervention for establishing norms and standards for sustainable design in the diagnostics sector. While TPPs generally define a broad range of characteristics regarding clinical need, analytical performance, clinical validity, infrastructure and human factors, and costs, environmental impact is less regularly considered in the writing process^3^. In an expert consensus from 2015/16, waste disposal was assigned low priority by experts in a panel discussion in the shaping of an AMR-related TPP^11^. In 2019 experts reviewed progress in the development of accurate, accessible and affordable rapid diagnostics tests and recommended that ‘environmental friendliness’ be added to the established, WHO endorsed, ASSURED criteria for assessing such devices^12^. TPPs are the best available tool for implementing such criteria and we recommend that considerations of environmental impact and waste disposal within TPPs should be given closer attention.

LFA kits are generally designed for single use. In kits containing multiple tests, some elements are reused on multiple tests within the kit, such as the IFU, quality card, sample holders, and the box itself. After use, the test and its associated components are meant to be disposed of, and LFA waste will generally end up in a municipal landfill or be incinerated. Thus, LFA testing generates a significant amount of waste which is not recovered in any way other than possible heat recovery from incineration. Material usage in such tests has never been rigorously investigated.

In this paper, we have collected and weighted the components of 21 different commercial COVID-19 lateral flow tests, with emergency use authorisations from the UK MHRA, EU, the US FDA, and the WHO EUL. We suggest this material usage mapping provides a comprehensive base for establishing future quantitative values for components in future TPPs.

## 2 Methods

We collected 21 different COVID-19 lateral flow test kits, with emergency use authorisations from the UK MHRA, EU, the US FDA, or the WHO EUL (Table 1). Despite significant efforts, it was easier to obtain tests available for personal use that had market authorisation in UK, than other tests. It was unsustainable to obtain many WHO-EUL tests as these were only available in large quantities from manufacturers and resellers: Artron, Wondfo, Flowflex for professional use had a minimum order range of 800-1000. The Wondfo and Flowflex self-tests available in UK do not have the same serial numbers as the WHO-EUL tests, but appear to originate from the same product line. Twelve samples were purchased in UK, five separate tests were purchased in France and Germany and another four samples were purchased in the US.

**Table 1:** Details of COVID-19 LFA kits collected.

| Test name | CE code | Ref code | Lot number | Country of purchase | Kit size | Number of individual tests weighed |
| --- | --- | --- | --- | --- | --- | --- |
| Everything genetic One Step Test for SARS-CoV-2 Antigen | CE1434 | CG20615 | LOTPSC225172W | UK | x1 | 3 |
| StepAhead One Step Test for SARS-CoV-2 Antigen | CE1434 | CG20615 | PSC220038W | UK | x1 | 2 |
| Flourecare SARS_CoV-2 Antigen Test Kit | CE1434 | MF-68 | n/a | FR | x1 | 1 |
| Medicspot Lateral Flow Antigen Travel Service | CE0123 | n/a | COV1110005 | UK | x1 | 3 |
| Flowflex SARS-CoV-2 Antigen Rapid Test | CE0123 | L031-118P5 | COV2065033 | UK | x5 | 3 |
| Orient Gene's Rapid Covid-19 (Antigen) Self-Test | CE0123 | GCCoV-502a-H70GE | 2201750 | UK | x7 | 3 |
| Hughes SARS-CoV-2 Antigen Rapid Test (Travel) | CE0123 | REF L031-118M3E | COV1070067 | UK | X1 | 1 |
| Getein One-Step-Test for SARS-CoV-2 Antigen | CE1434 | REF CG206155 | PSC220040W | UK | x5 | 3 |
| Wondfo 2019-CoV Antigen Test | CE0123 | W634P0024 | W63410603 | UK | x1 | 3 |
| Wondfo 2019-nCoV Antigen Test | CE0123 | W364P0028 | W63410604 | UK | X2 | 3 |
| Boson Rapid SARS-CoV-2 Antigen Test Card | CE0123 | 1N40C5-4 | 220112091 | UK | x5 | 3 |
| Panbio COVID-19 Ag RTD Nasopharyngeal | n/a | 41FK10 | 41ADH256A | DE | x25 | 3 |
| BinaxNow COVID-19 Antigen Self Test | n/a | REF 195-160 | 213253 | USA | x2 | 3 |
| iHealth COVID-19 Antigen Rapid Test | n/a | GTIN 20856362005894 | 221CO20130 | USA | x2 | 2 |
| QuickVue At-HomeOTC COVID-19 Test | n/a | 20402 | F41691 | USA | x2 | 2 |
| Ecotest Covid-19 Antigen Nasal Testkit | CE1434 | COV-535002H5 | I2112213 | DE | x5 | 3 |
| NHS Test&Trace COVID-19 Self-Test (RAT) | n/a | TK2193 | X2201712 | UK | x7 | 3 |
| Hotgen Coronavirus (2019-nCov)-Antigen Test | CE0123 | HGCG134S0101 | W2022030400 | DE | x1 | 1 |
| Safecare COVID-19 Antigen RTK | CE1434 | COV Ag-6012H | COV22072001 | DE | x1 | 1 |
| Newgene COVID-19 Antigen Detection Kit | CE1434 | COVID-19-NG21 | 20220718-01 | DE | x1 | 1 |
| Ellume COVID-19 home test | n/a | I-SRS-C-01 | QB02S-H | USA | X1 | 1 |

We took tests apart manually, first simply opening all packaging. A scalpel was used to dissemble the LFA cassettes and extract the nitrocellulose strip or other similarly integrated components. The dismantling process is illustrated by a photograph in *Supplementary Figure S1*. An individual component was considered a component that cannot be further taken apart, for example, because it is made of a single block of plastics. These individual components were weighed using a Fisherbrand PS-60 precision scale for weights below 60g, and an Ohaus CL series scale for weights above 60g. Categories and sub-categories of components of interest were created. Where possible, at least three identical tests were weighted and total weight and individual component weights were averaged. For boxes with multiple tests, the weight of a single test was calculated by dividing the total of shared component weights by the number of tests. A series of photographs illustrating the content of each individual kits in the collection is available in *Supplementary Figure S2*.

## 3 Results

### General observations and total weight of LFA kits

A list of all identified test components and sub-components can be found in Table 2. Briefly, all test kits in the collection contained a card or plastic-based outer packaging solution, instructions, swab pack (swab to obtain nasal or nasopharyngeal samples packed in sterile paper and plastic packaging), cassette pack (usually consisting of an aluminium foil pouch containing a nitrocellulose strip encased in hard plastic cassette, and a desiccant packet); a reagent pack (usually simply consisting of a pre-filled reagent tube). Beyond these key components, some test kits also include additional contraptions, for example, a test tube holder, or rack, when the outer packaging itself is not serving as a tube holder. Many, but not all kits, include one biohazard or waste zip-lock bag per test to gather all used test components for disposal, or a thin certification paper slip. Test kits can also contain additional interior packaging such as large soft plastic zip-lock bags.

| Component categories | Sub-component categories | Further division of sub-component categories |
| --- | --- | --- |
| A. Outer packaging |  |  |
|  | A.1 Box |  |
|  | A.2 Outer foil pouch |  |
|  | A.3 Plastic Wrapping |  |
|  | A.4 Additional cardboard packaging |  |
| B. Instructions |  |  |
|  | B.1 IFUs |  |
|  | B.2 Additional information |  |
| C. Certification slip |  |  |
| D. Biosafety/ Waste bag |  |  |
| E. Swab pack |  |  |
|  | E.1 Swab |  |
|  | E.2 Packaging part 1 (paper) |  |
|  | E.3 Packaging part 2 (plastic) |  |
| F. Reagent pack |  |  |
|  | F.1 Tube |  |
|  | F.2 Tube foil lid |  |
|  | F.3 Tube plug |  |
|  | F.4 Dropper (if separate from tube) |  |
|  | F.5 Screw-on cap |  |
|  | F.6 Blister pack |  |
|  | F.7 Reagent |  |
|  | F.8 Additional Bags/Packaging |  |
|  | F.9 Additional Bags/Packaging (foil) |  |
|  | F.10 Additional Bags/Packaging<br>Rubber band |  |
|  | F.11 Dessicant |  |
|  | F.12. Reagent bottle |  |
| G. Cassette pack |  |  |
|  | G.1 Packaging |  |
|  | G.2 Dessicant |  |
|  | G.3 Test cassette |  |
|  |  | G.3.1 Lateral flow strip |
|  |  | G.3.2 Outer casing top |
|  |  | G.3.3 Outer casing bottom |
|  |  | G.3.4 Battery |
|  |  | G.3.5 on/off button |
|  |  | G.3.6 plastic LFA strip tray top |
|  |  | G.3.7 plastic LFA strip tray bottom |
|  |  | G.3.8 Analyzer PCB |
| H. Tube Rack |  |  |
| I. Control Swabs |  |  |

The total weight of the kits, normalised by the number of individual tests, ranged from 15 to 85 g per individual tests, with a mean of 33 g ± 19.6 g (*Figure 1A*). These statistics illustrate both the disparity of designs between manufacturers and the possibility to decrease individual kit weights.

**Figure 1.**
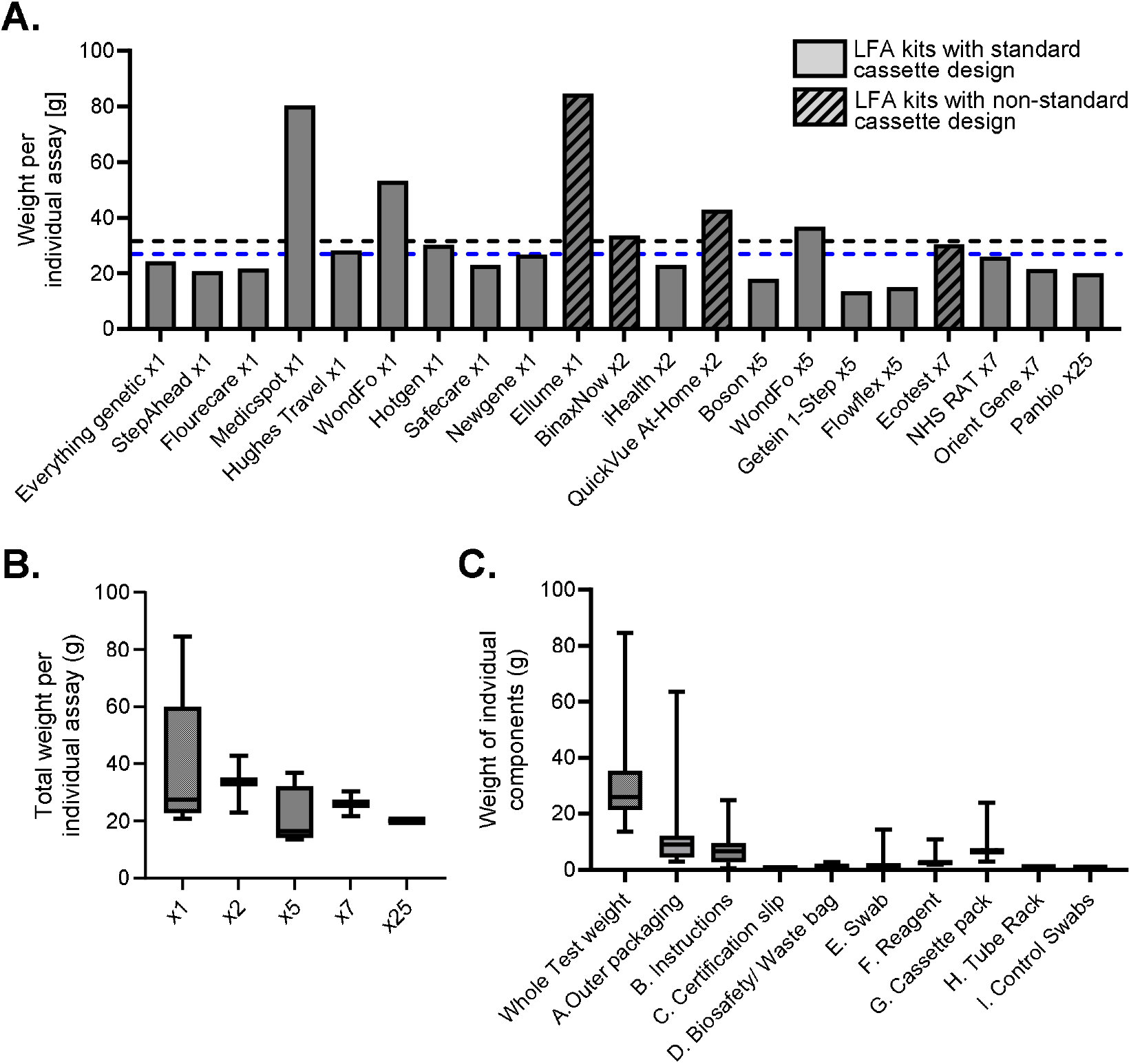
Total weight of LFA tests by A) brand B) number of tests per kits (single ‘x1’, or multi-test kits of two, five, seven or twenty-five tests). C) Average whole tests weight and average weight of individual components across the whole collection. In the box plots, the whiskers show the minimum and maximum. The horizontal bar in the box shows the median.

The heaviest kit, Medicspot, stands out because it is a single Flowflex test repackaged in a larger heavier cardboard box alongside a small card containing information on how to register the test to obtain a travel certificate.

The collection includes single test (‘x1’, N=10) kits and multi-test in two-test (‘x2’, N=3), five-test(‘x5’, N=4), seven-test(‘x7’, N=3) and twenty-five-test (‘x25’, N=1). One test, the Wondfo kit, is represented in both single-test and two-test kits. The weight of a single Flowflex (inside the Medicspot packaging), can also be compared to the Flowflex five-test kit. In both case, the multi-test kits required less overall material per test. Packing five tests instead of one, results in 30%, and 39% less overall weight, respectively in the Wondfo and Flowflex kits. Whilst an overall similar trend can be observed by comparing the average test weights of all single (39 g), and two-, five-, seven-, and twenty-five-pack kits (respectively 33 g, 21 g, 26 g and 20 g), the variations between the kits are too large for this trend to be statistically significant (Anova test, p-value = 0.5) (*Figure 1.B*).

The weighting of the individual components of the kits enables further analysis about which components have the most influence on the total weight of the kit, and where these variations come from. The three heaviest components on average are the outer packaging (mean 11.5 g ± 13 g), cassette pack (mean 7.7 g ± 4.2 g) as described above, and instructions (mean 7 g ± 5.9 g) (*Figure 1.C*). The outer packaging drives the largest weight variations seen in the total kit weights. However, this is largely driven by the Medicspot kit, which is a Flowflex single-pack kit nested, as previously described, in a large outer packaging. If considering this kit an outlier, the component with the largest variations is the instructions, followed by the cassette pack.

### Cassette designs and cassette weights

Seventeen out of the twenty-one COVID-19 LFA tests feature a nitrocellulose strip in a shallow hard plastic cassette casing, with a highly consistent design, usually around 2 cm wide, 6-9 cm long, and around 0.5 cm tall. Out of these seventeen similar-looking designs, they are still a 22 % of deviation in weight (Outer casing top (G3.2) and outer casing bottom (G3.3) combined), with an average of 4.1 g ± 0.9 g and a range from 2.5 g to 6.5 g (*Figure 2*).

**Figure 2.**
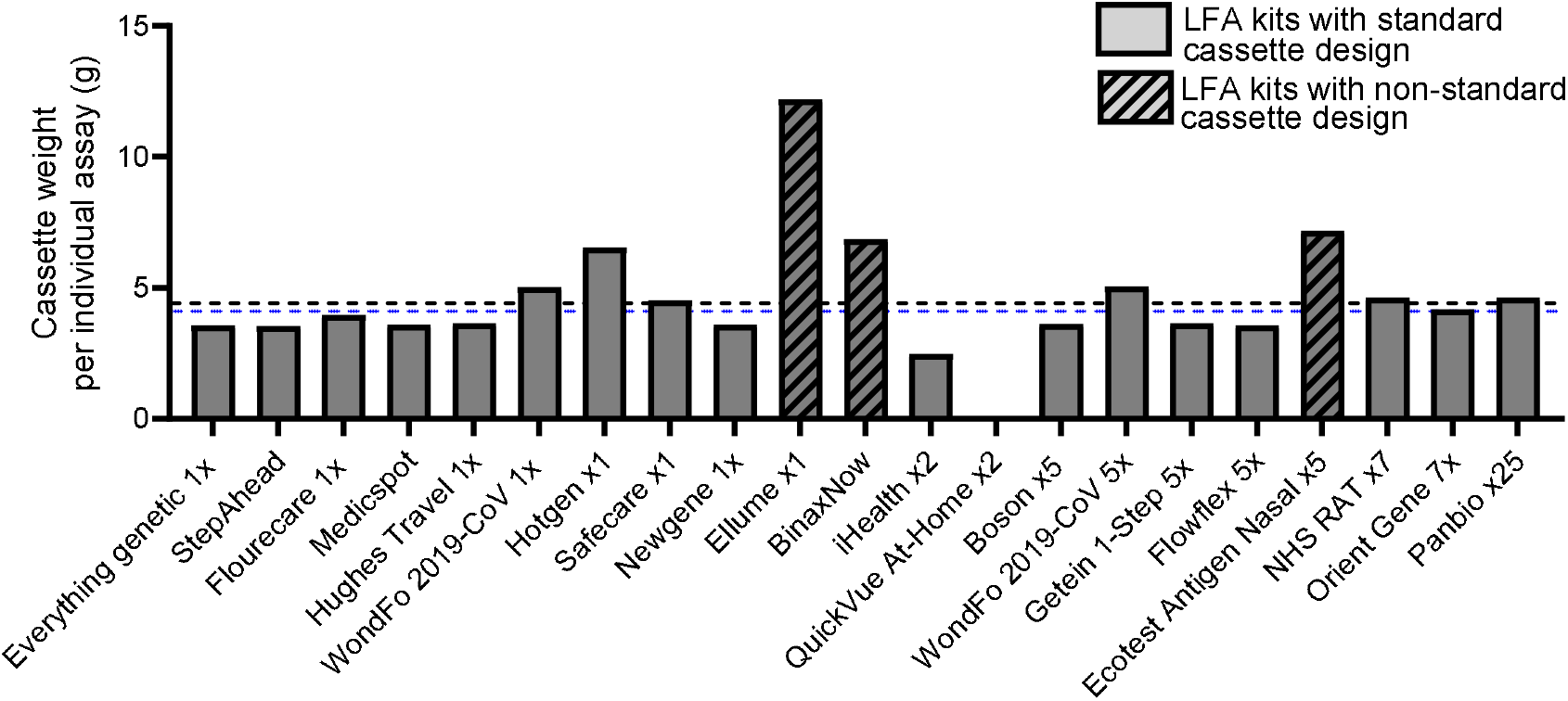
Weight of the hard plastic cassette casing sub-component (combined G3.2 and G3.3) in individual tests. Black dash line (mean all standard and non-standard tests at 4.57 g) Blue dash line (mean all standard and non-standard tests at 4.10 g)

Four of the kits in this collection have a non-standard cassette design for encasing the nitrocellulose strip. The US FDA EUA-approved QuickVue test does not use a test cassette at all. The extraction tube, already pre-filled with reagent, is used to conduct the test: after taking the sample and mixing it in the tube, the swab is removed and the LFA test strip, which is stored on its own without casing or desiccant in a foil pouch, is dipped into the tube. The BinaxNow uses a cardboard cassette (no hard plastic) for the lateral flow test strip, and the reagent is dripped onto the swab with the sample inside the cardboard cassette. Otherwise, it is packaged and set up like a cassette test. Also included, is a digital battery-powered COVID-19 test produced by Ellume, which includes a large hard-plastic casing weighing 12.2 g. Anecdotally, the Ellume cassette was the hardest to take apart from all the cassettes investigated. This raises the question of how many customers will be taking the battery button from the device before disposing of the test. Additionally, we included the Ecotest, a test available for purchase in Germany at the time of collection, which takes the form of a pen-shaped test that combines a swab, cassette, and extraction tube into one pen-shaped device. The equivalent cassette element weighs 7.2 g. A photograph of all extraction tubes is available in Supplementary Figure S.3.

### Swab designs and swab weights

Swabs are usually purchased from a small range of suppliers rather than manufactured alongside the test, hence this is a component that has the lowest variations amongst the kits. The weights provided describe the swab only (component E.1). The average weight of swabs in the 16 standard design kits is 0.43 g, with a range from 0.26 g to 0.74 g. The main difference between swabs is the length and thickness of the swab shaft, which is reflected in the overall weight (Figure 4). Additionally, swab lengths were found to have an impact on the overall packaging size – longer swabs require larger boxes and larger waste bags. Large differences were found in the non-standard kit, with the Ellume swab, and QuickVue test weighting respectively 9.8 g, and 1.51 g, and featuring among the top two heaviest swabs. A photograph of all extraction tubes is available in Supplementary Figure S.4.

**Figure 4.**
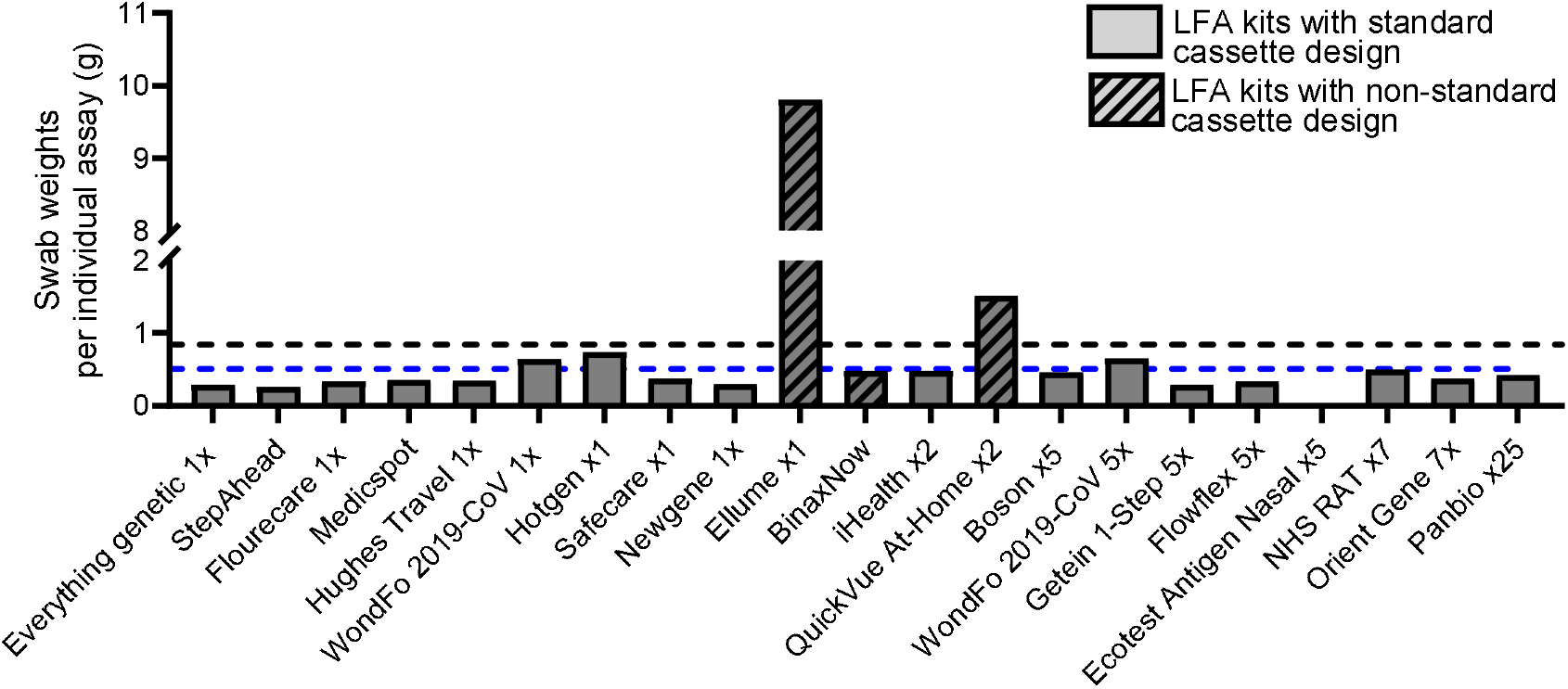
Swab component (E) weight in individual tests. Black dash line (mean of all standard and non-standard tests at 0.87 g) Blue dash line (mean all standard tests at 0.43 g)

**Figure 5.**
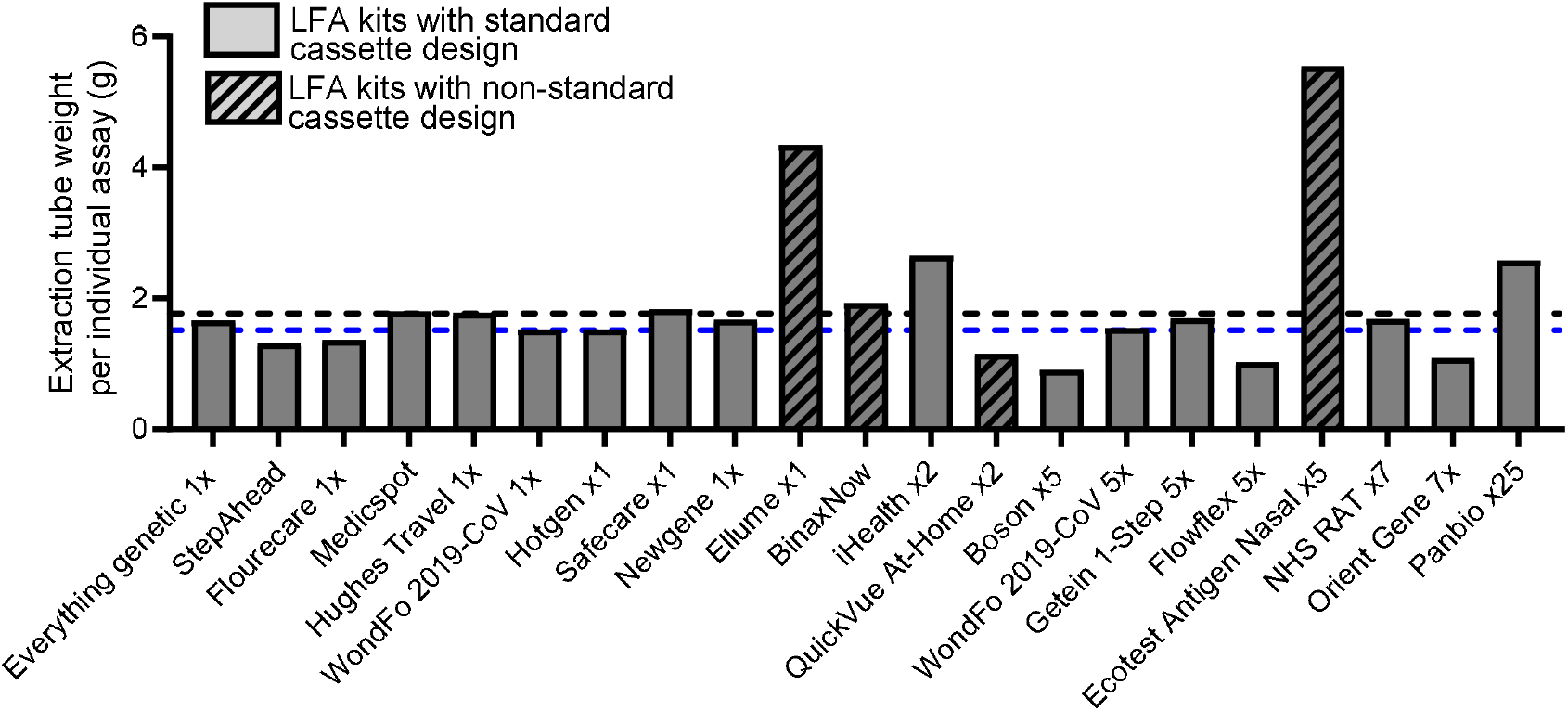
Weights of extraction tube component (F1) in individual tests. Black dash line (mean all standard and non-standard tests at 1.97 g) Blue dash line (mean all standard tests at 1.63 g)

### Extraction tube designs and weights

The biggest conceptual difference between test kits lies in the packaging of the reagent and the design of the extraction tube. The majority of tests use the extraction tube to also store the reagent liquid. The Boson kit, which uses a separate reagent blister and extraction tube, and the Panbio test, which provides 9 ml reagent in a reagent bottle for 25 tests, are exceptions (Photographs is Suppl.Fig S5). Extraction tubes are closed either with foil lids or screw-on caps, which has little effect on the component weights overall. The weights provided in this section describe the empty tube only (component F.1). The average weight of reagent tubes in the test kits was 1.8 g if only tubes were included, 1.85 g if tubes and reagent blisters were counted jointly, not including the weight of any packaging materials. The heaviest reagent tube design weighs in at 5.55 g (Ecotest), the lightest at 1.3 g (StepAhead). In the standard design kits, the heaviest reagent and extraction tube solution used a tube, a screw-on cap, and an additional screw-on cap or plug to secure the liquid.

### Packaging weight fraction in different kits

Packaging design and decisions are highly impactful on the overall weight of the kit, and thus its environmental footprint. All packaging related components (components A, B, C, D and H) were grouped together, and their weights compared to the core test components (components E,F,G, I). Large variations in the distribution of packaging weight across different components of the selected LFA tests were observed (Figure 6). In 14 out of 21 kits, all related packaging was observed to represent a larger weight fraction than the test cassette and other ancillary parts necessary to conduct the assay, with packaging making up on average 57% of the whole test kit weight, with large variations observed between 34% and 89%. Abbott (manufacturer of the BinaxNow and Panbio tests) addressed packaging in an article on their website, announcing they had reduced packaging weight and volume for both test lines by removing internal plastic trays and reducing package size in line with their 2030 Sustainability Plan^13^. The QuickVue test has the lightest test of the collection, yet one of the highest total weights due to a significant amount of packaging materials including a large plastic tray. This contrast indicates that the manufacturer may have been trying to reduce its costs on the cassette weight (the most expensive plastic element in the LFA test), without adhering to environmentally sustainable design principals. It is also possible that manufacturers are hesitant in selling small boxes to customers, which could fail to be seen on pharmacies, para-pharmacies or supermarket shelves, or appear of lower value to the customer.

**Figure 6.**
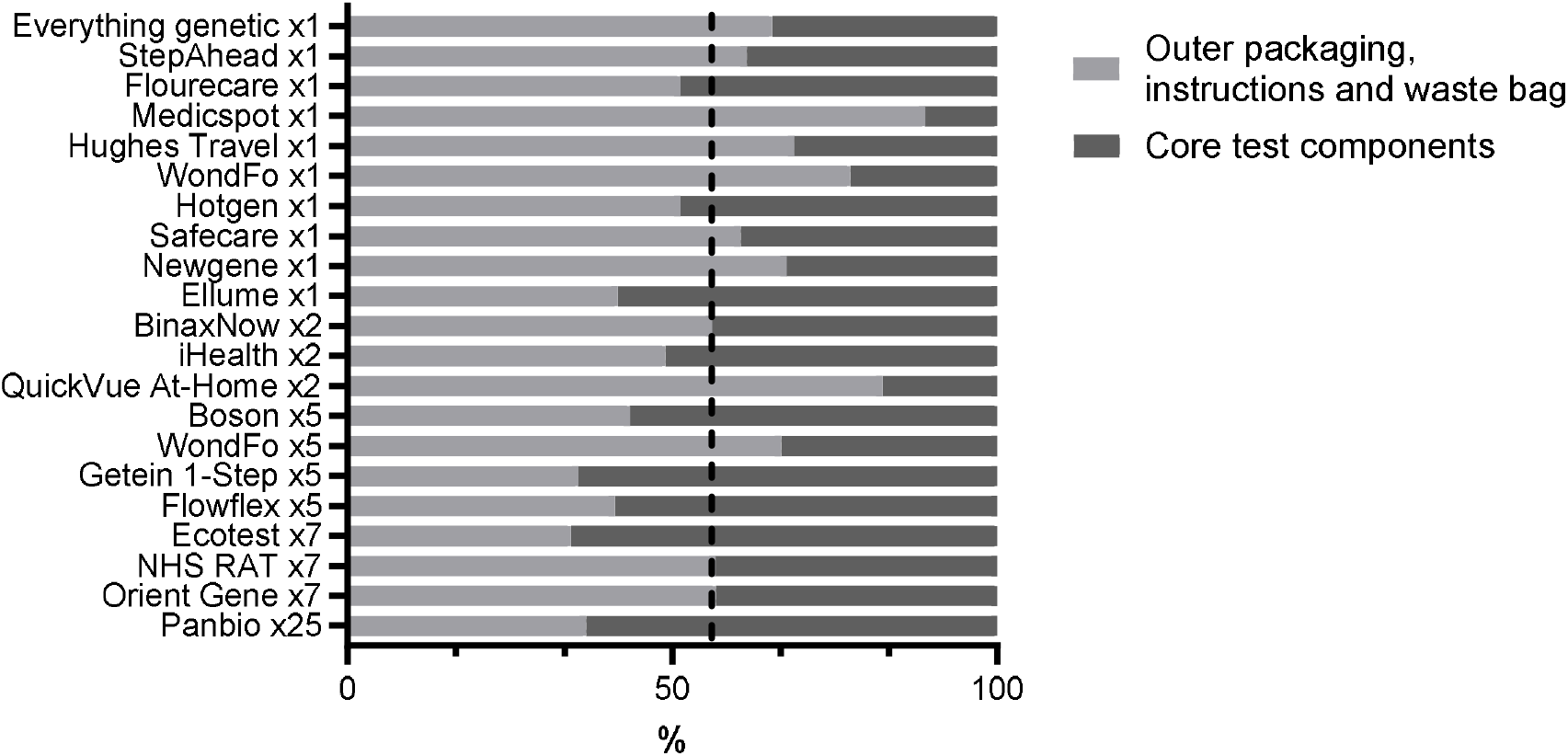
Comparison of the weight of packaging-related component and the weight of core test components

Peripheral to packaging, but also of note, are the different weights of the instruction booklets. Instructions vary in length between brands. This depends partially on the number of languages required – a test sold in the UK generally only includes English instructions, a test sold in the US also usually includes a Spanish instruction booklet. WHO EUL-listed tests usually come with English-language instructions, though some manufacturers provide additional translations in a variety of languages.

## 3. Discussion

We investigated a range of lateral flow assay (LFA) test designs commercially available for at-home COVID-19 testing in the UK, USA, and EU, comparing component weights across designs to highlight differences in material use even amongst almost identically-shaped cassette tests. The results give an unprecedented comparative view of design and weight fractions in a wide range of commercial COVID-19 tests. These results give us a quantitative analysis of representative current industrial practice in terms of LFA component design and weight.

The collection included seventeen standard designs and four non-standard designs. The collection also included single-test and multiple-test kits. Overall, the multi-test kits were shown to have a limited impact on the overall weight of the individual tests, but large variations between kits limit the validity of the analysis. When comparing identical kits in single and five-test options, the five-test kits were found to bring a reduction of 35% in the overall kit weight.

Amongst the standard designs, the component or sub-components cassettes, swabs and tubes were still found to display significant variations with CVs of respectively, 20%, 36% and 28%. We hypothesize that some manufacturers may decrease the weights of these components for financial reasons, not environmental ones.

The non-standard designs were found to be on average 1.8 times heavier than the standard designs. The analysis of the non-standard design kits against the standard design kits revealed that non-standard designs add an average of 60% on the cassette sub-component. However, individual components might vary significantly between kits. Two of the non-standard kits feature the two lightest cassettes: the BinaxNow and Quickvue, whilst the two other non-standard kits feature the two heaviest cassettes, the Ellume cassette has a 197% added weight with respect to the average of the standard kit, and the Ecotest adds 75%.

This illustrates that (i) there is scope for much lighter cassette designs and (ii) without policy guidance, innovative designs can go either way in terms of material usage and environmental sustainability.

Light core test components did not necessarily translate into overall lighter kits, notably, the Quickvue test had the lightest test (a bare nitrocellulose strip weighing 0.29 g) but the kit was packaged using a large plastic tray, and as a result, the whole package contained the most heaviest packaging (per single test) of all weighted kits. We hypothesise again that manufacturers do not generally reduce material usage because of environmental concerns but for economic reasons. They might subsequently increase material usage in cheaper components, such as packaging in order to increase visibility on the shelves of supermarkets or pharmacies.

These market-average weights reveal both the possibilities and pitfalls in material usage. Although the overall weight of LFA kits rarely exceeds 30 g, these devices are produced out of virgin plastics in billions every year, generating a significant added amount of medical waste, and production is expected to increase year on year with new types of tests appearing on the market, such as nucleic-acid based LFAs^14,15^.

We propose that market-average weights could form the basis for quantitative recommendations for the sustainability sections of TPP, which have until now been qualitative only, and far from ambitious. The average weight of several of the components, including the cassette, swabs, and packaging, could be used to set minimum and optimal weight thresholds. We propose to target specific elements, in line with current industry practice, namely the weight of the cassette, swab reagent and extraction tube. The suggested terminology could be “*weights no higher than the market average of x*” for individual components, and “*kit packaging weight (divided by individual test for multiple test kits) should not exceed x % of total kit weight*” (Table). The measures could effectively drive industry practice towards inconspicuous and light designs, preventing the development of overly material-heavy devices, favouring the market towards the use of lighter materials (e.g. natural cellulose) and the ultimate abandonment of plastics.

## 4. Conclusions

Over 2 billion LFA test kits are produced every year, adding tens of thousands of tonnes of used materials to the growing global volume of medical waste. TPPs are powerful tools that influence choices made by medical manufacturers in terms of form factors, design, functionality, and performance. However, they are seldom used to influence choices in terms of environmental sustainability.

Here a representative sample of LFA industry practice was analysed by selecting, disassembling, and weighing all individual components of 21 different commercial lateral flow assays with emergency use authorisations from the UK MHRA, EU, the US FDA, or the WHO EUL.

This study demonstrates that the market is not homogenous. Although most manufacturers favor a particular design of cassette, significant differences remain in the weight of various components, including cassettes, swabs, tubes, and importantly, packaging choices. The quantitative data collated from twenty representative professional or home COVID-19 LFA tests form a clear picture of the baseline in terms of the quantity of material used. These market-average weight measurements highlight the possibility of reducing overall material usage, including plastic usage in LFA test components.

Policymakers, such as the WHO, FIND, and others, are encouraged to make use of this dataset to derive target weights of individual components or full test and develop quantitative environmental sustainability targets in future Target Product Profile documents for lateral flow assays.

In the medical device sector, and in particular in point-of-care product development, Target Product Profiles are influential documents. However, they rarely include environmental sustainability considerations. The data produced in this study can help close this gap and provide quantitative data for target values not to exceed when designing and producing a new product, thus addressing the growing environmental burden of health care by reducing reliance on virgin, petrochemical-based plastics in point-of-care diagnostics.

## Supporting information

LFA Databas

## Data Availability

All data produced in the present work are contained in the manuscript.

## Acknowledgements

We are grateful to the Edinburgh Earth Initiative Earth Fellows programme for supporting this research.

## Funding

This project has received funding from the European Research Council (ERC) under the European Union’s Horizon 2020 research and innovation programme under grant agreement No 715450.

## Competing interests

none

## Notes

### Competing Interest Statement

The authors have declared no competing interest.

### Funding Statement

This project has received funding from the European Research Council (ERC) under the European Union Horizon 2020 research and innovation programme under grant agreement No 715450.

