## Supplementary Material for "Material usage in commercial lateral flow assay kits"

**Supplementary information**

**Figure S.1. Two photographs illustrating the process of taking the kit apart and weighing whole kits or kit components**

**[To comply with MedrXiv policy, these photographs were removed from the submission, readers should contact the corresponding author to request access to these materials]**

**Figure S.2. Series of photographs illustrating the sub-components of each test in the collection**

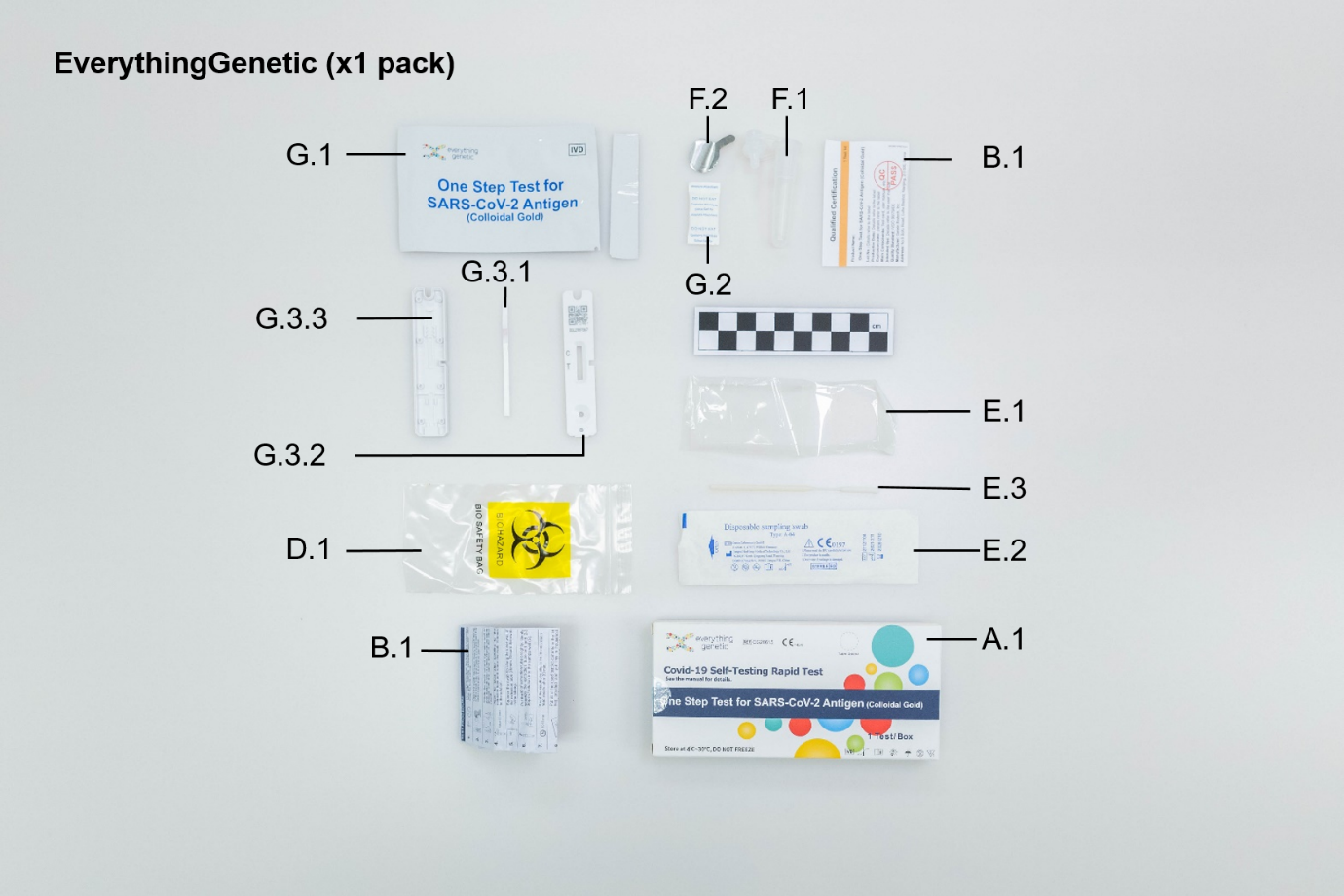

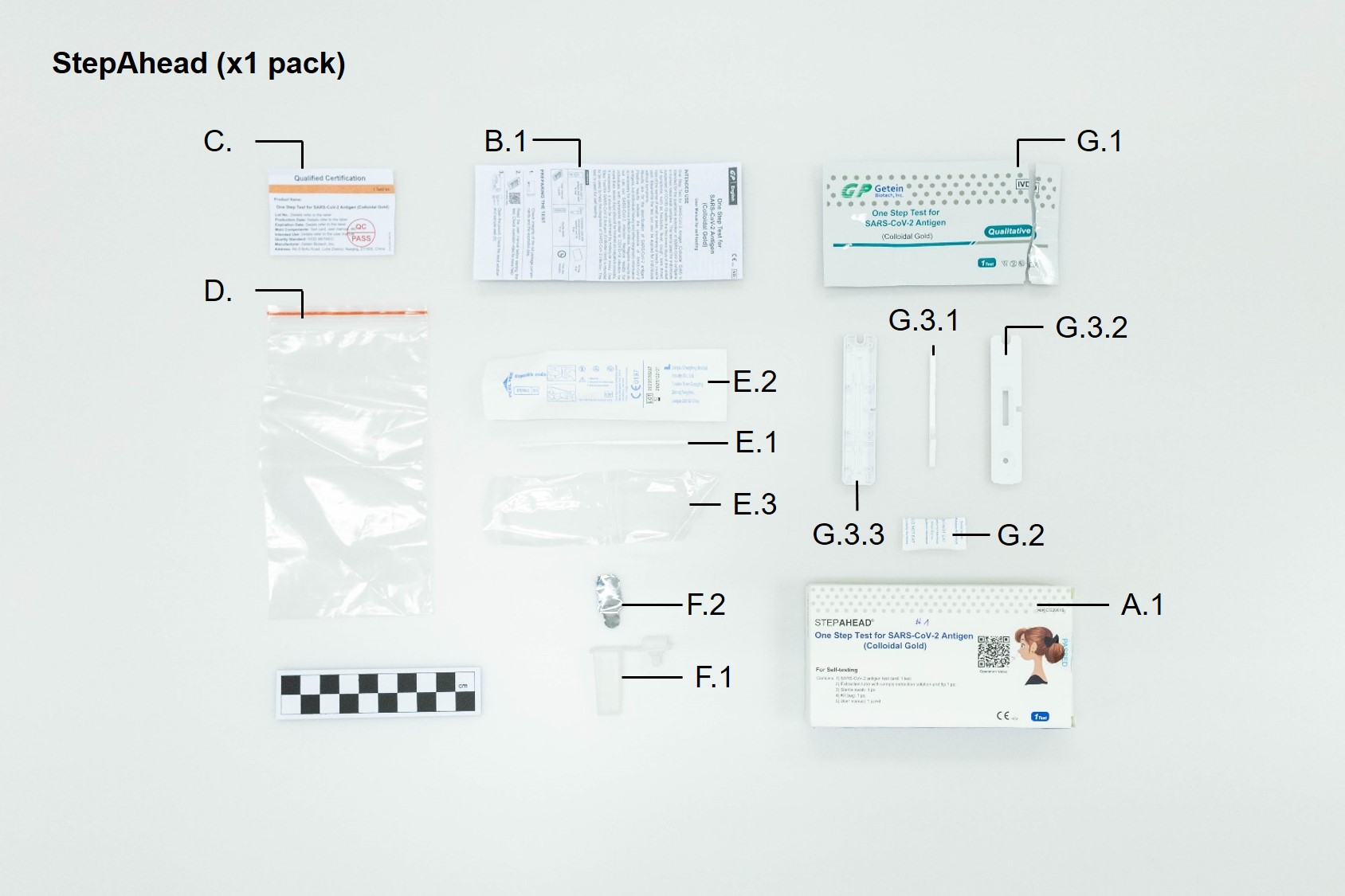

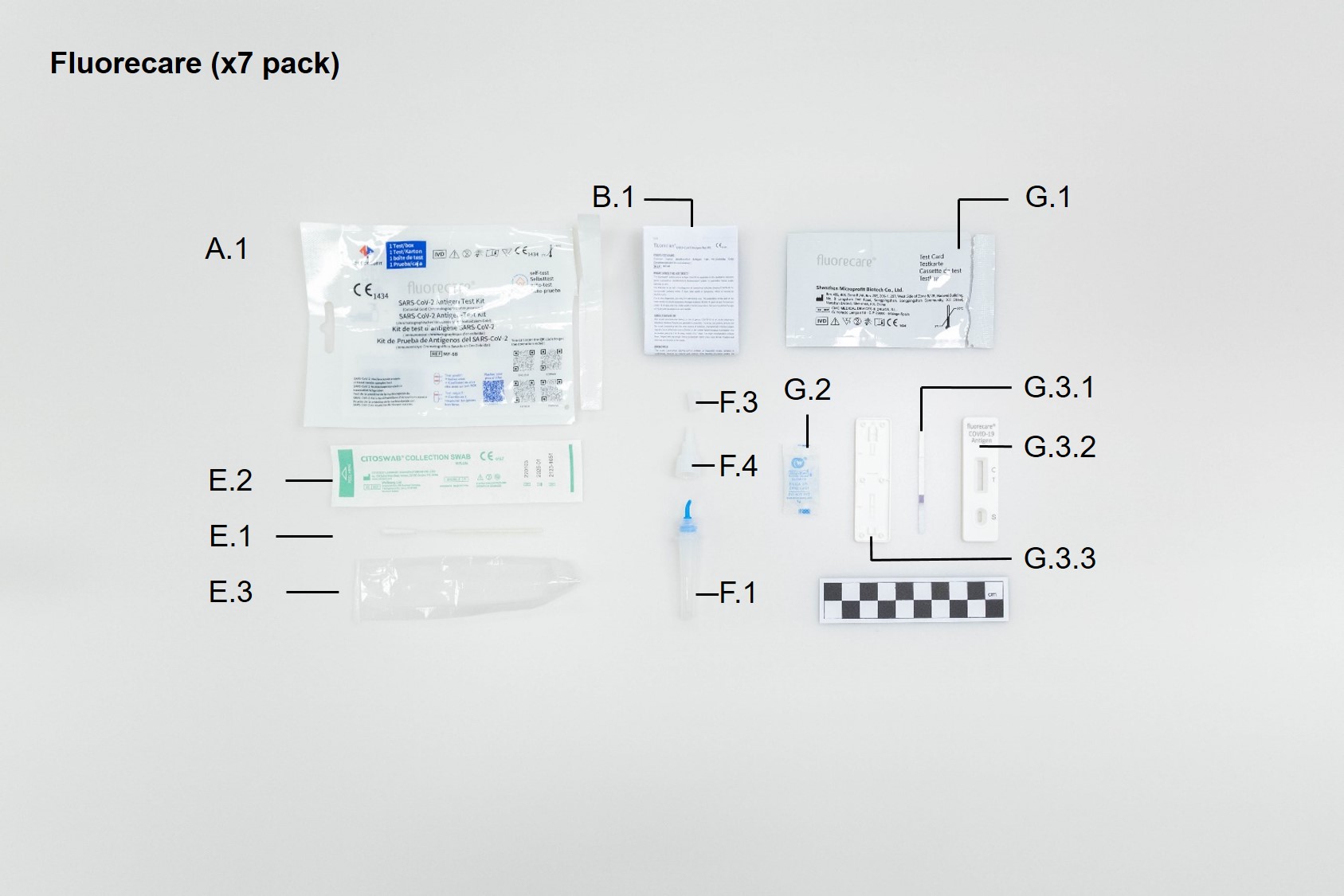

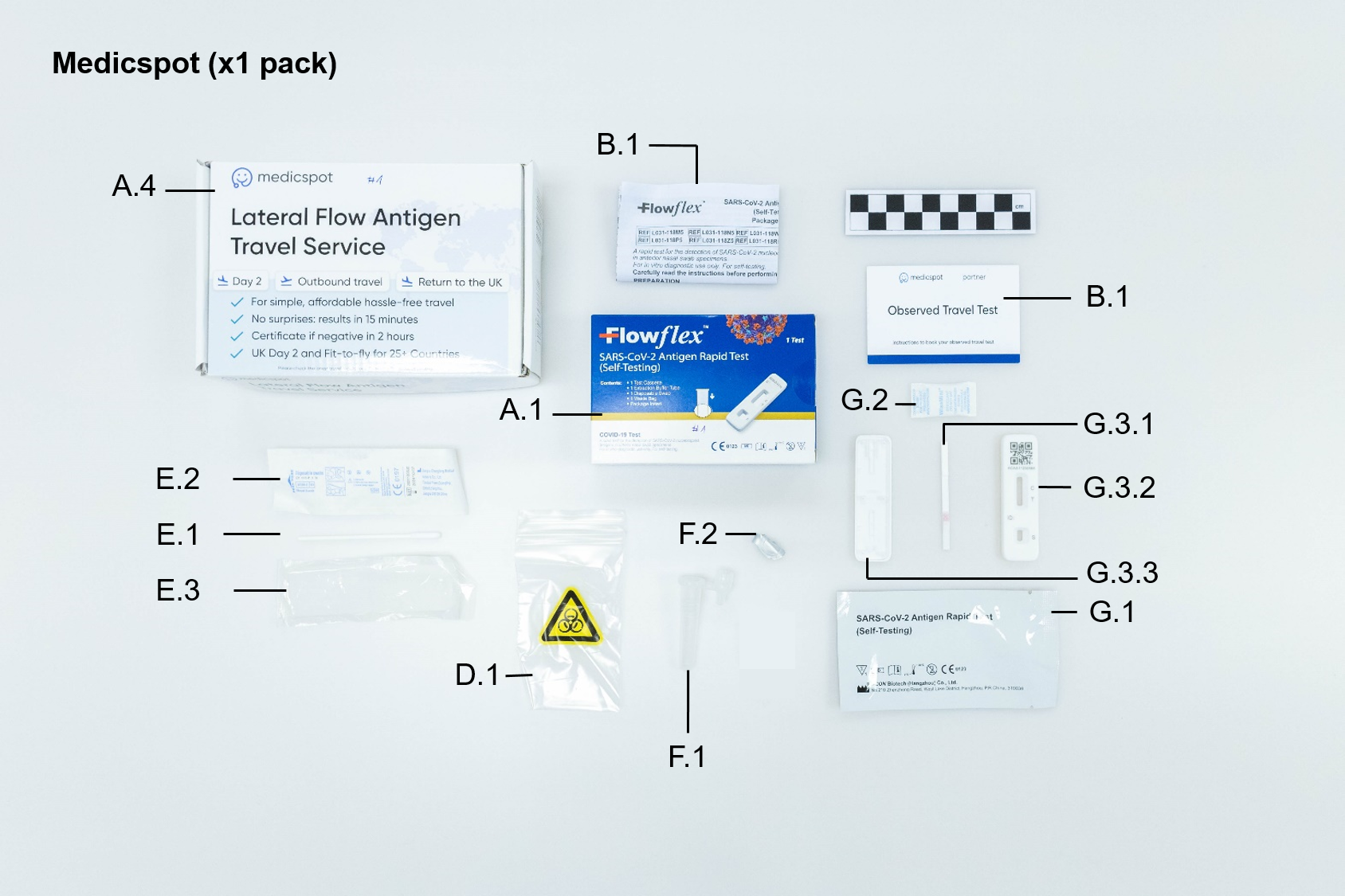

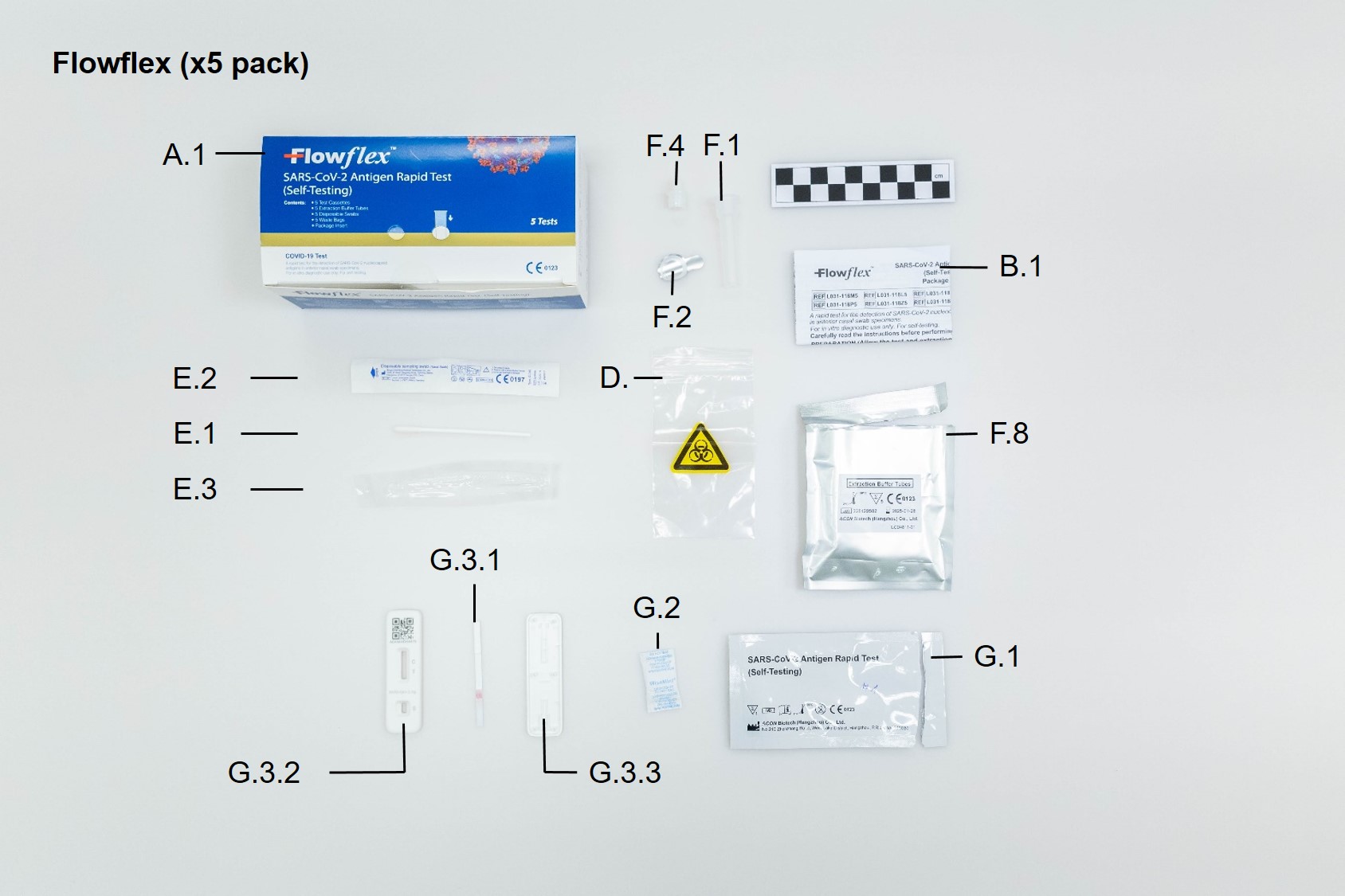

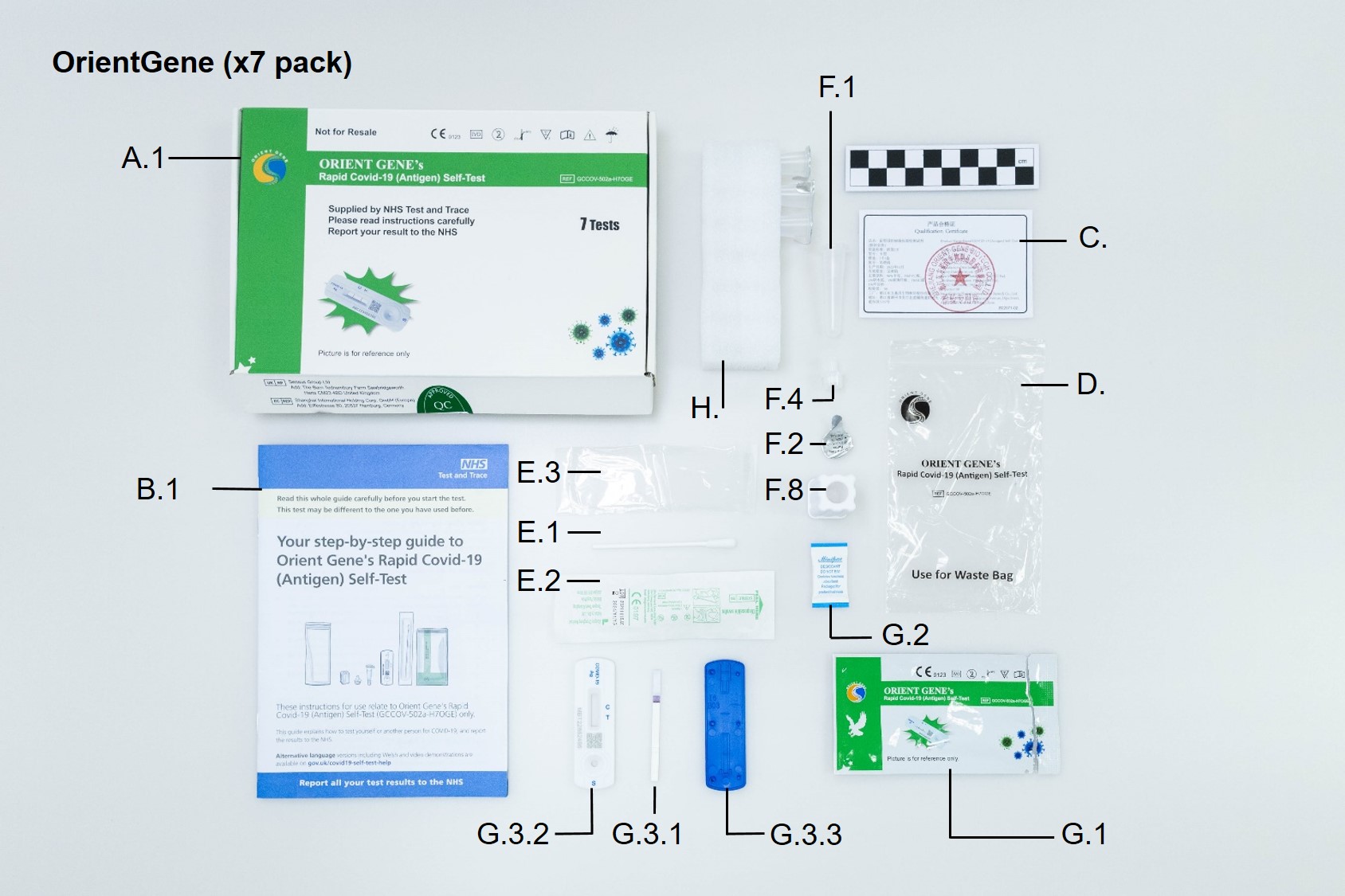

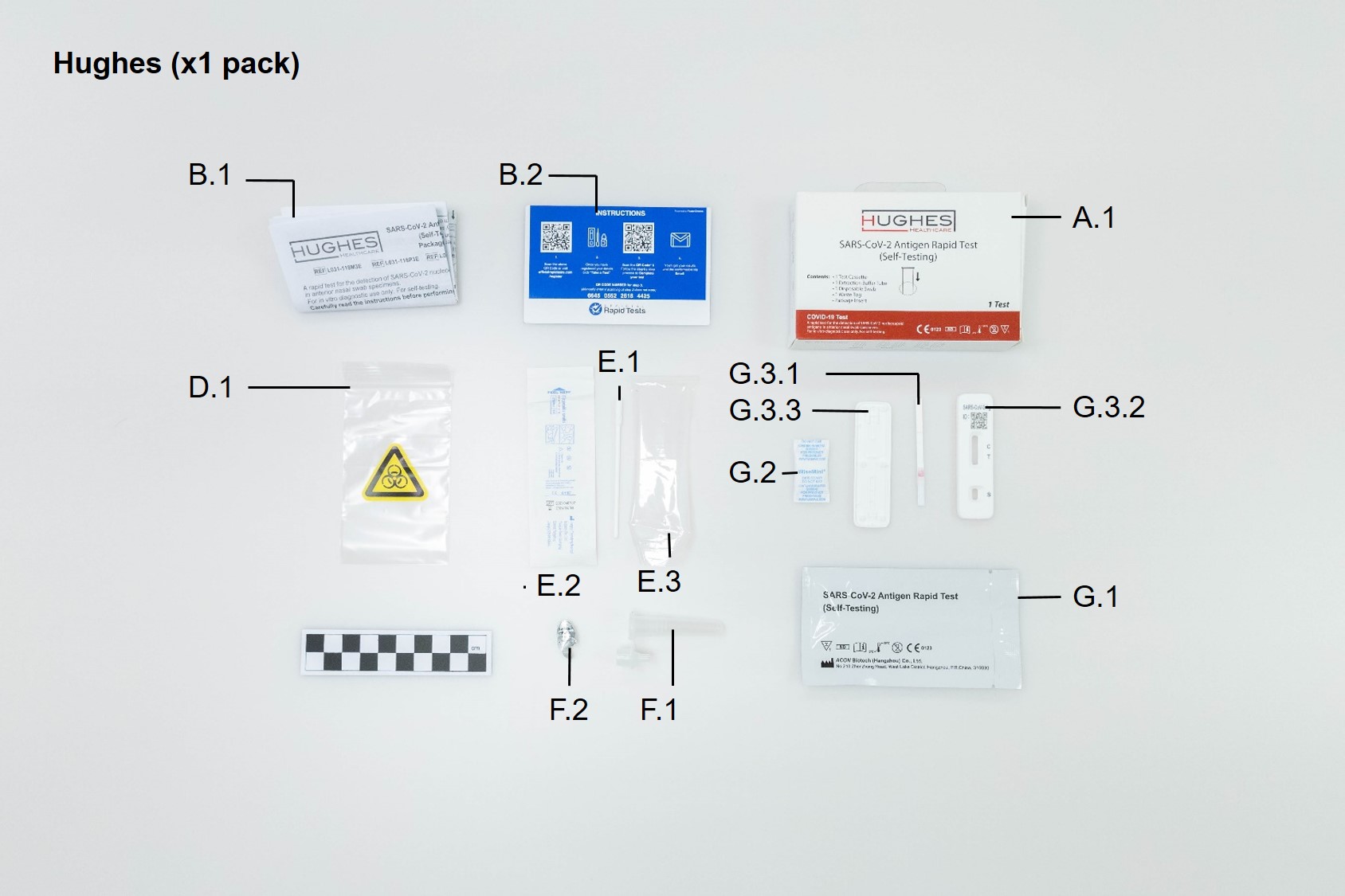

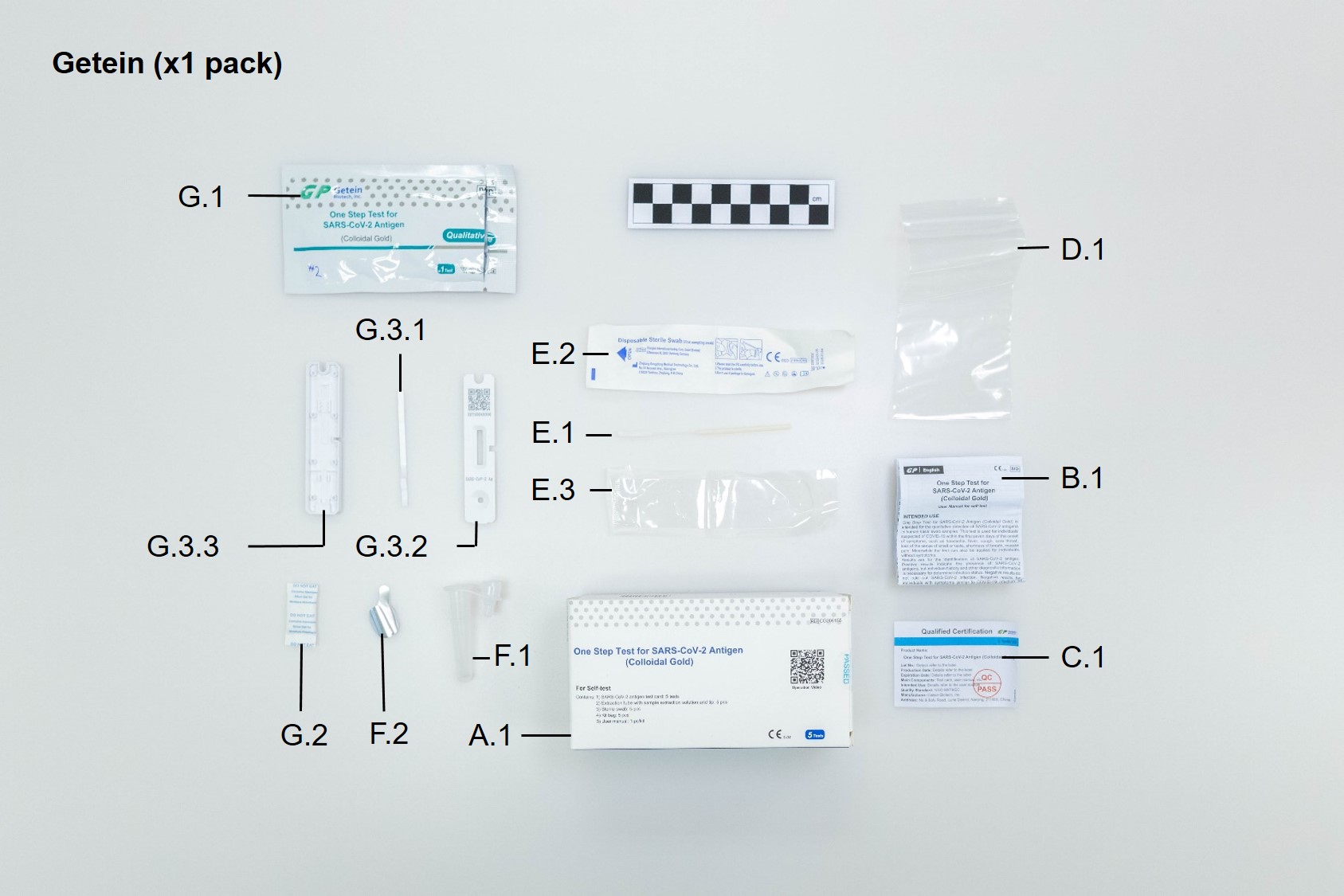

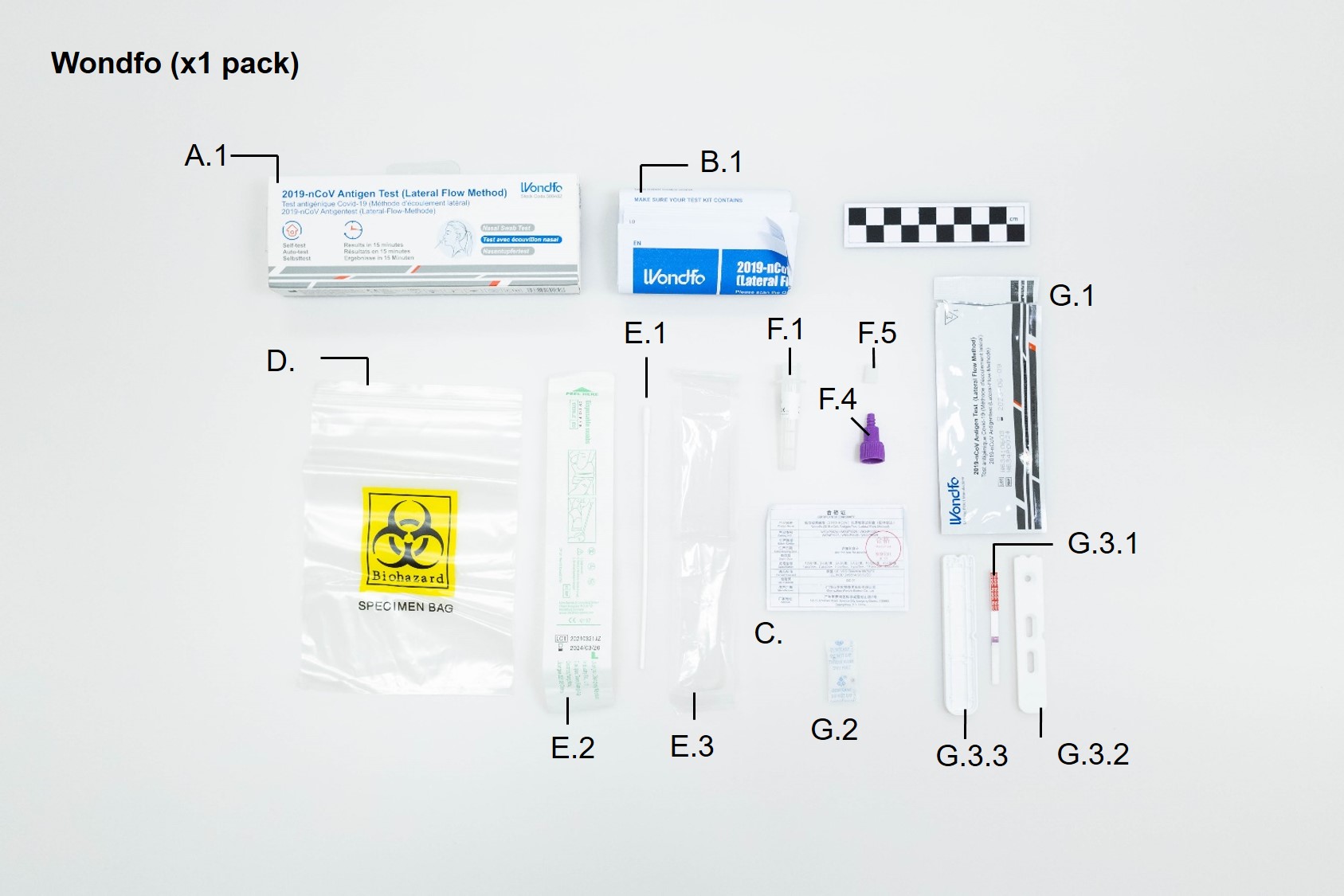

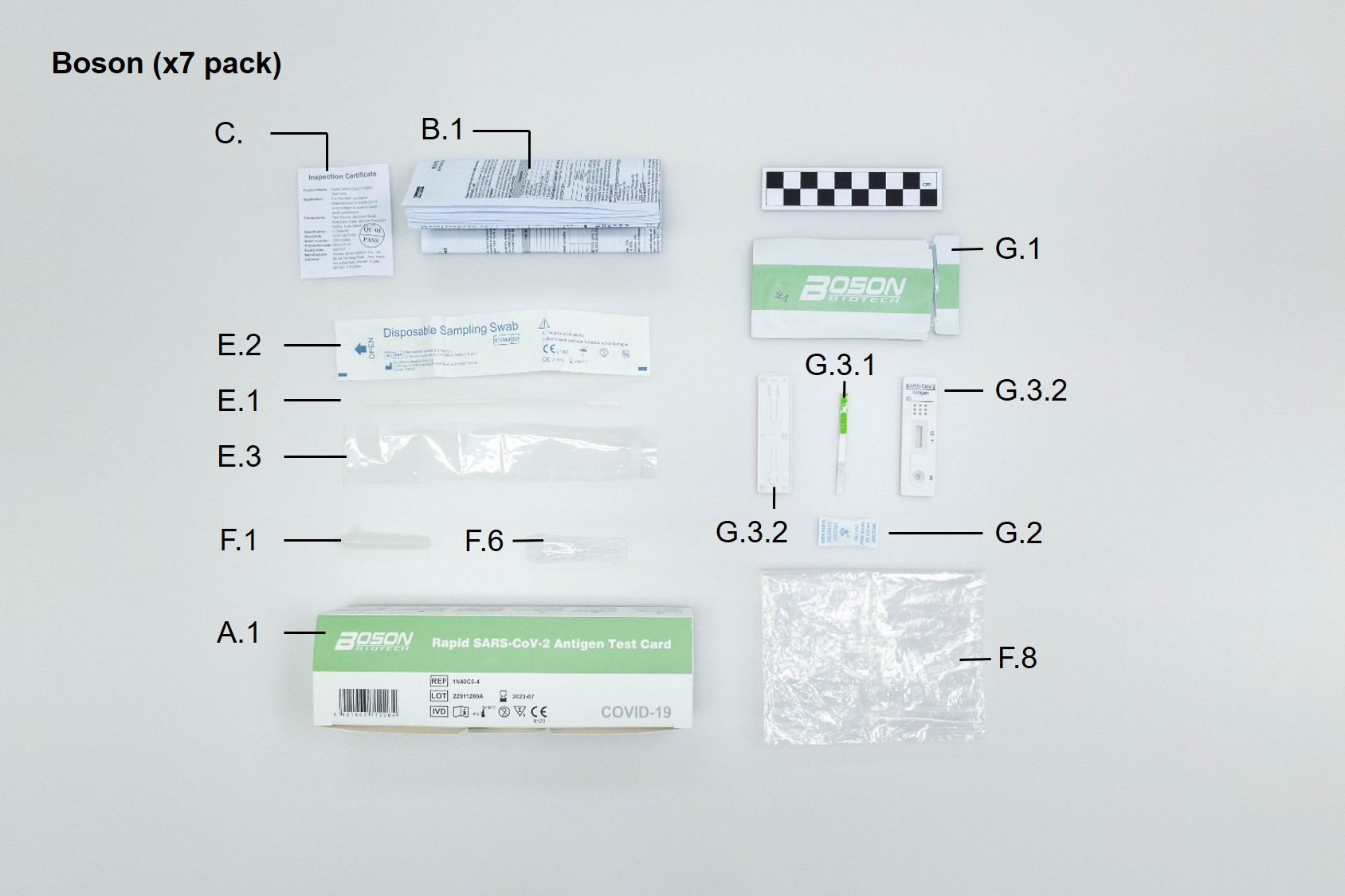

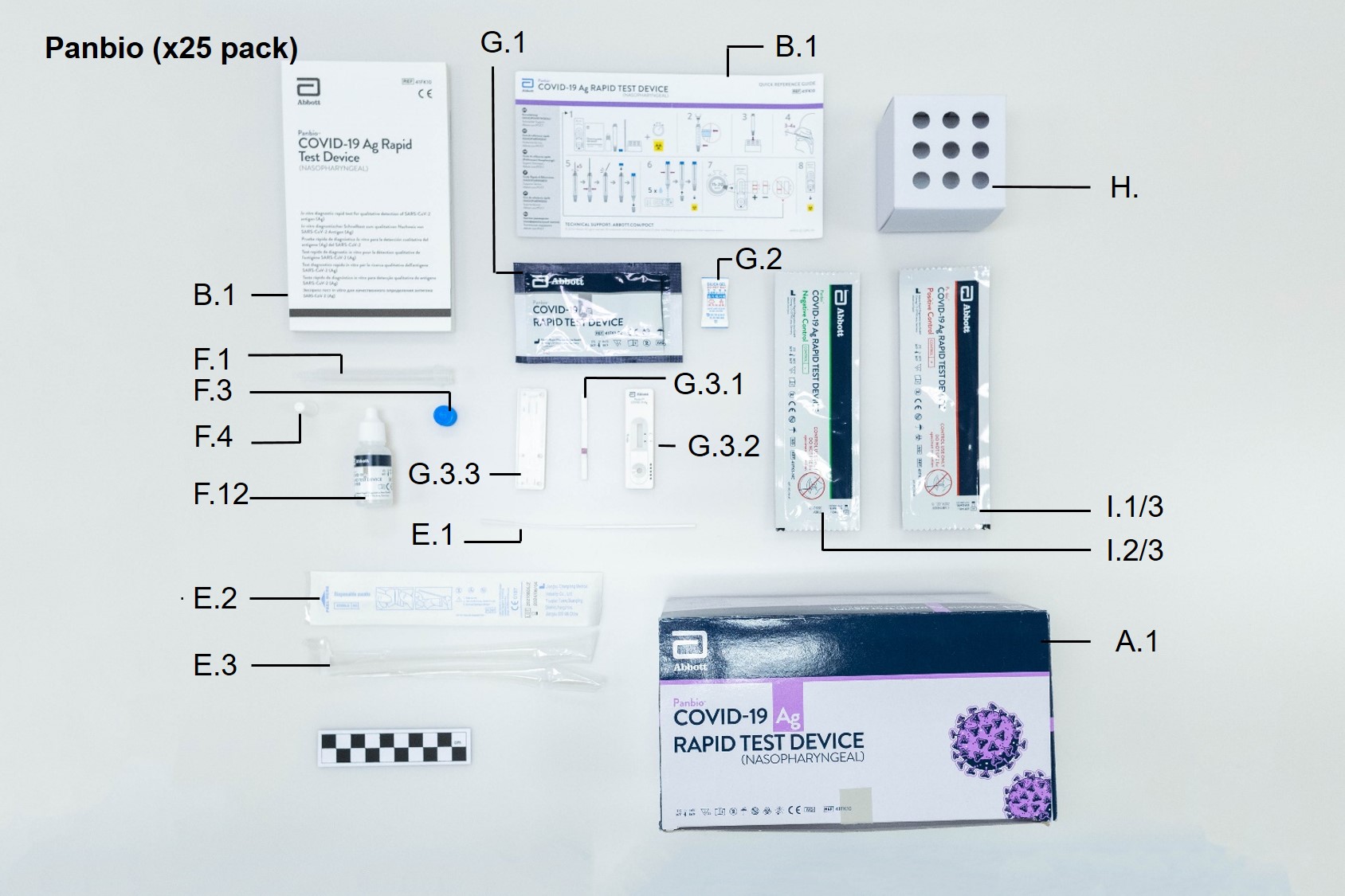

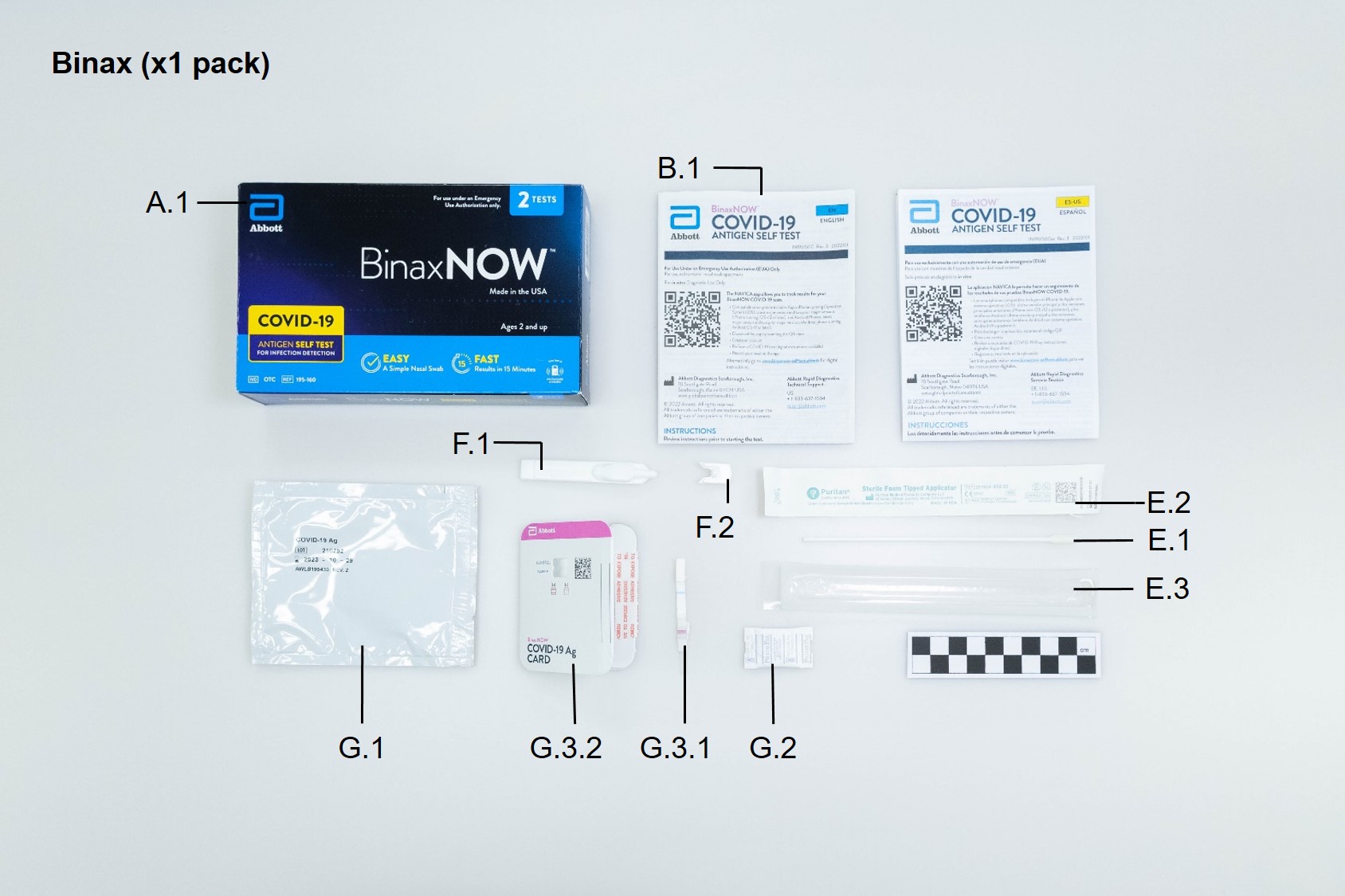

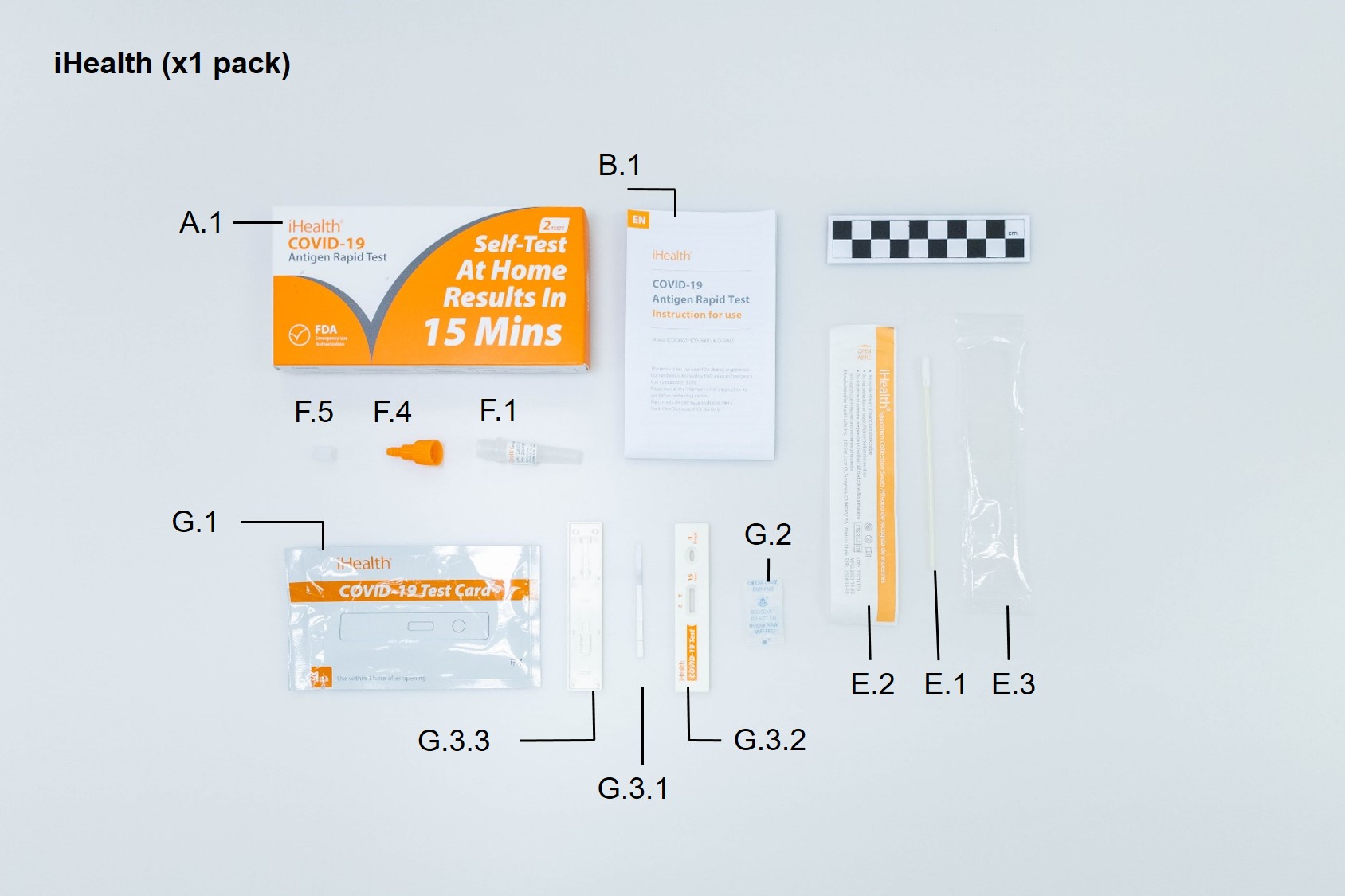

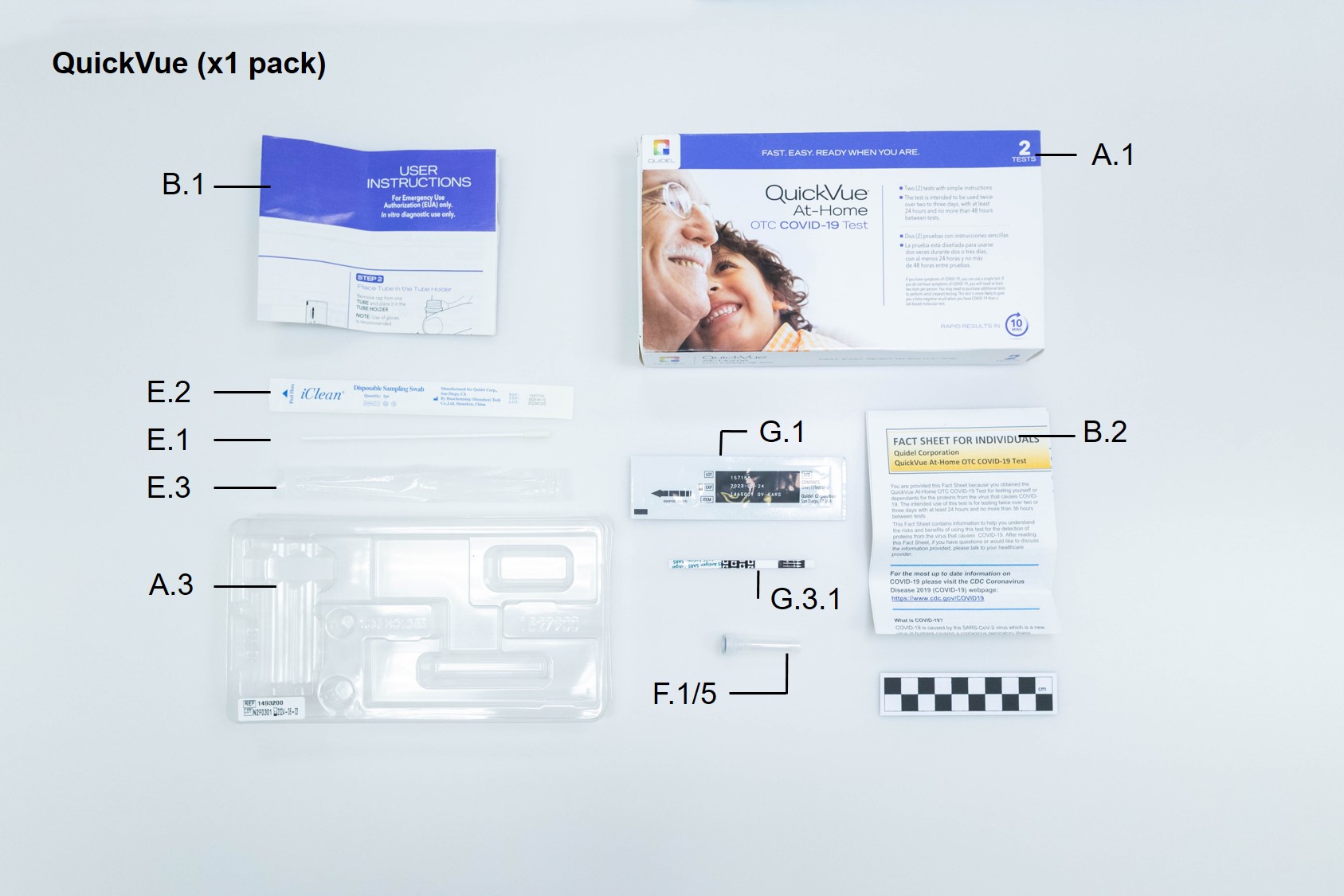

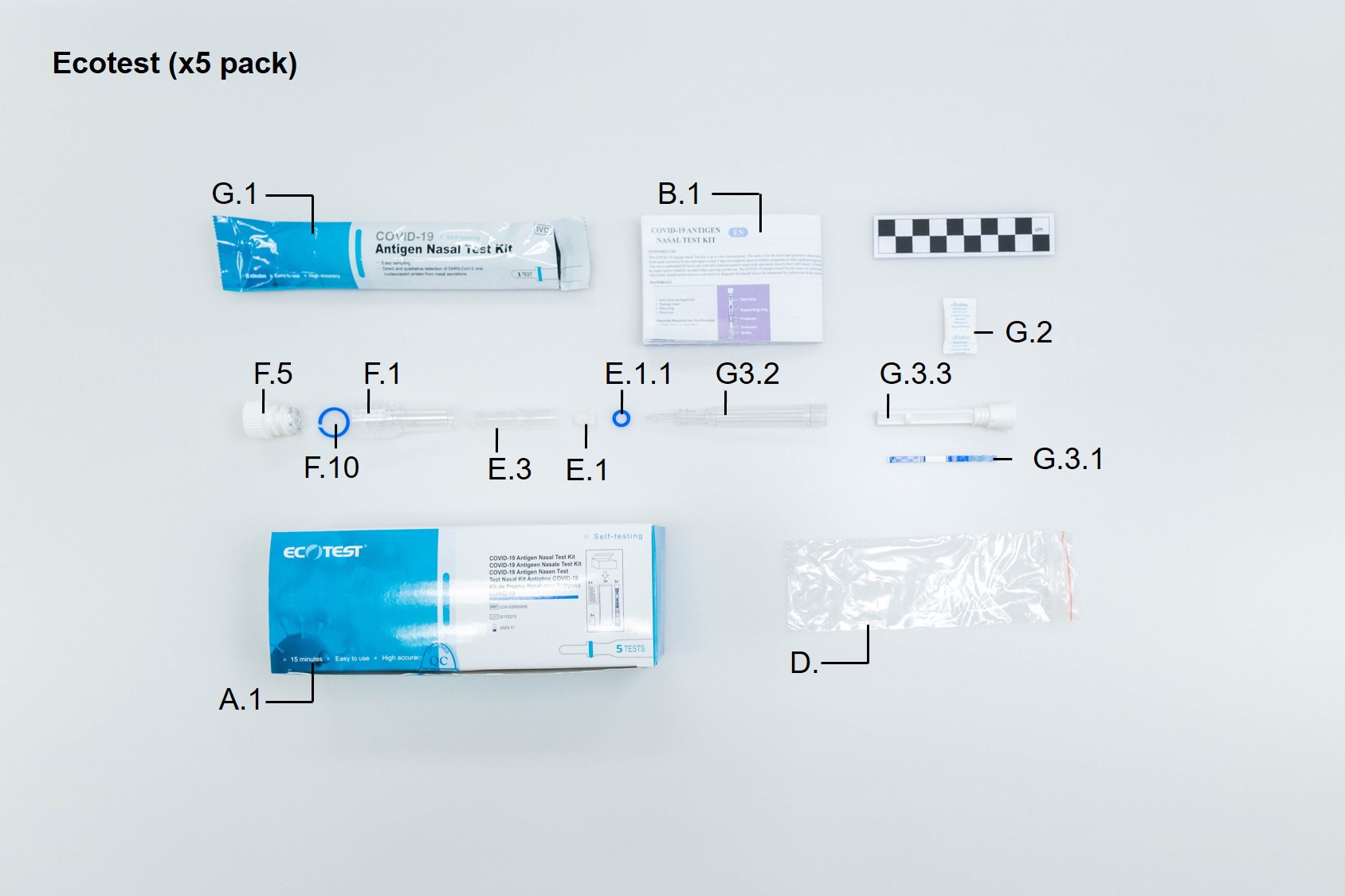

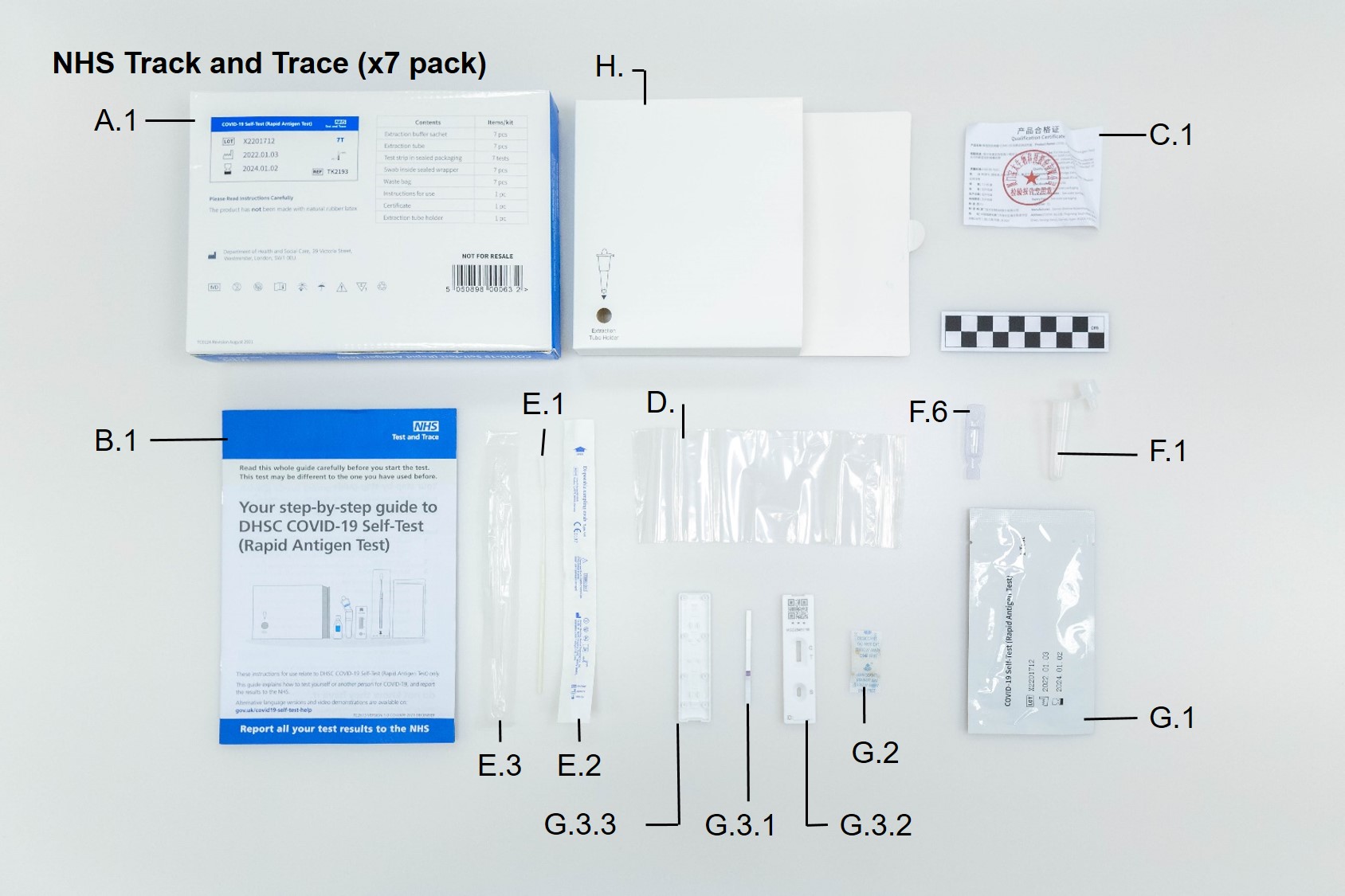

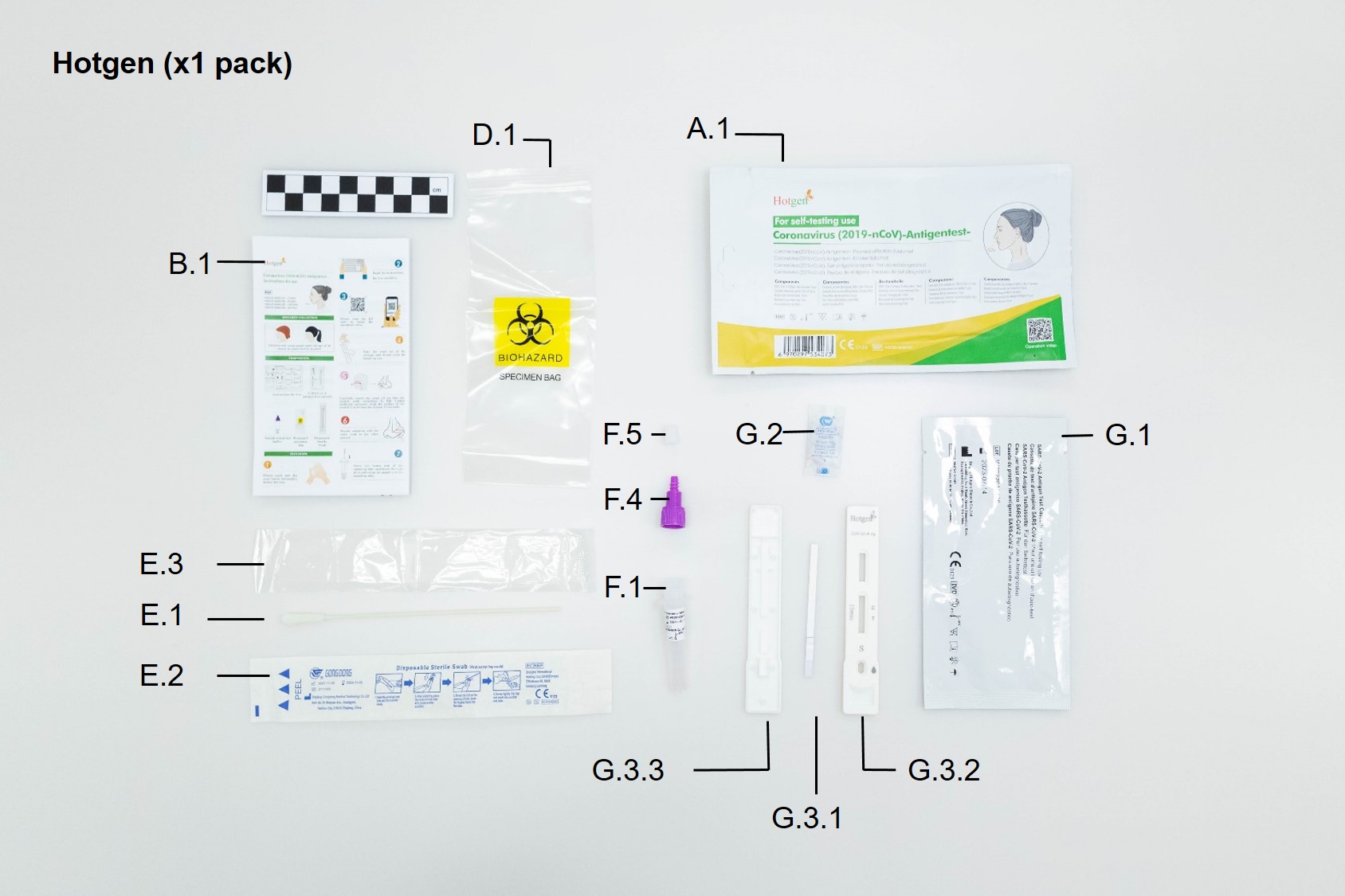

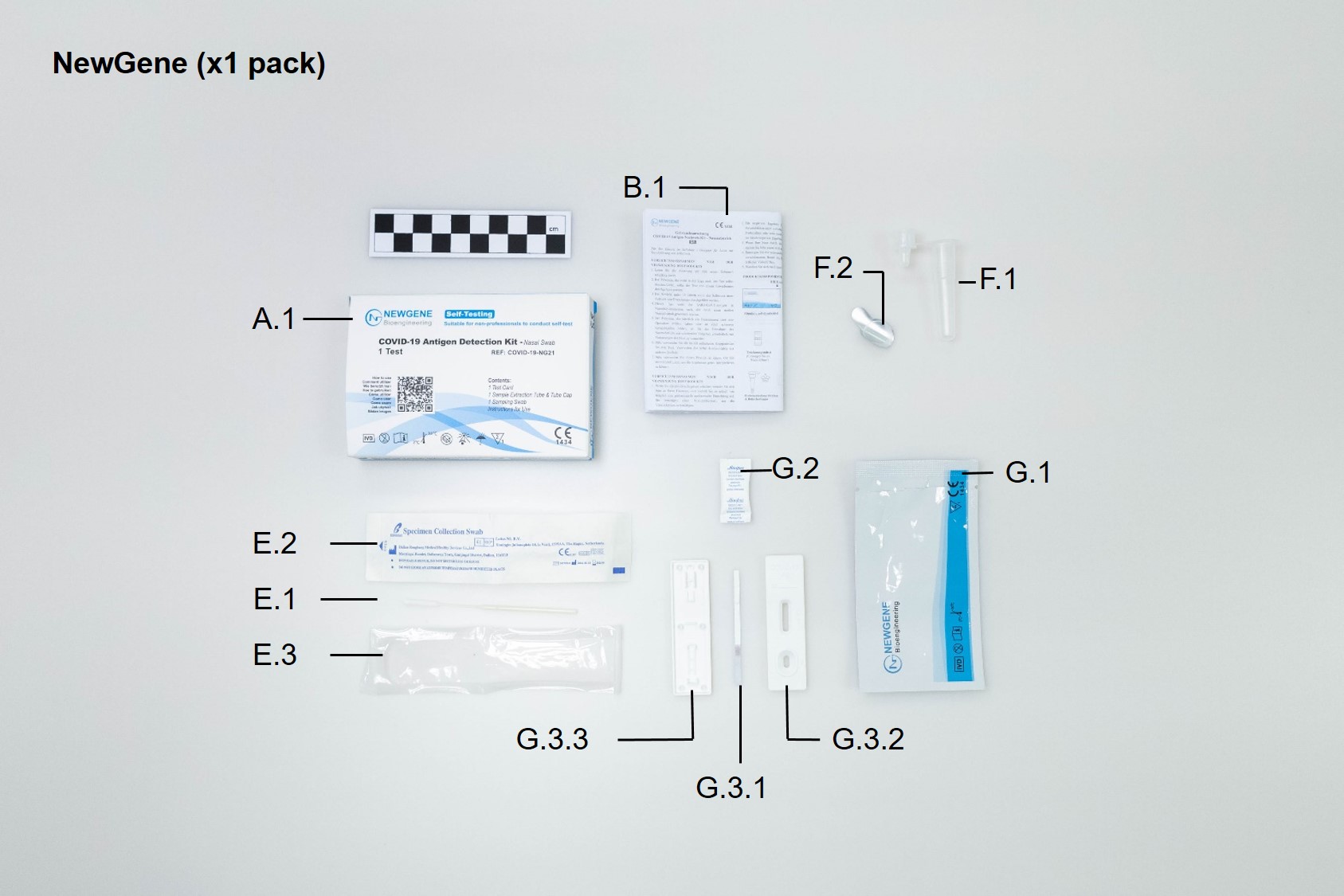

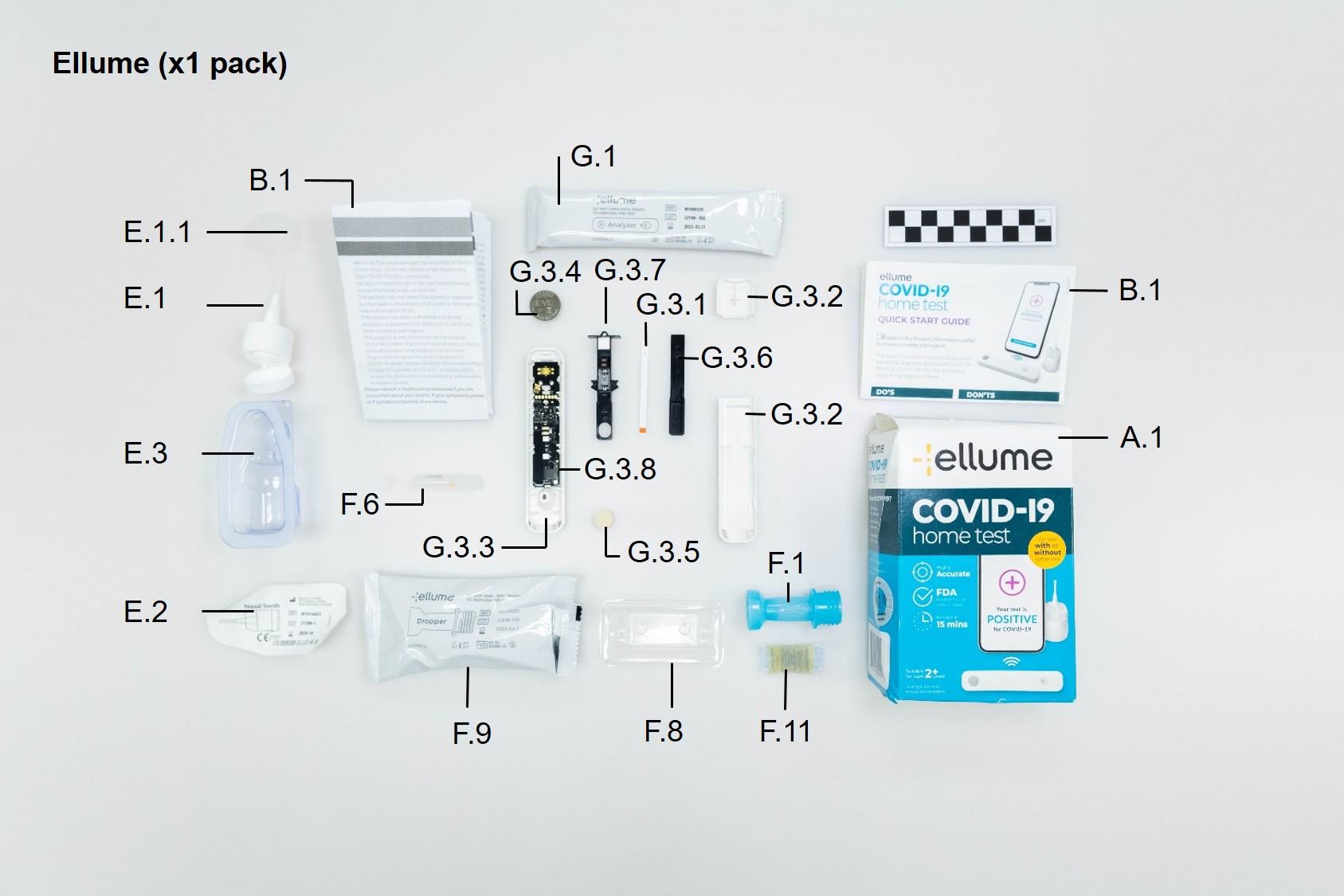

**Figure S.3. Photograph of all cassette designs in the collection**

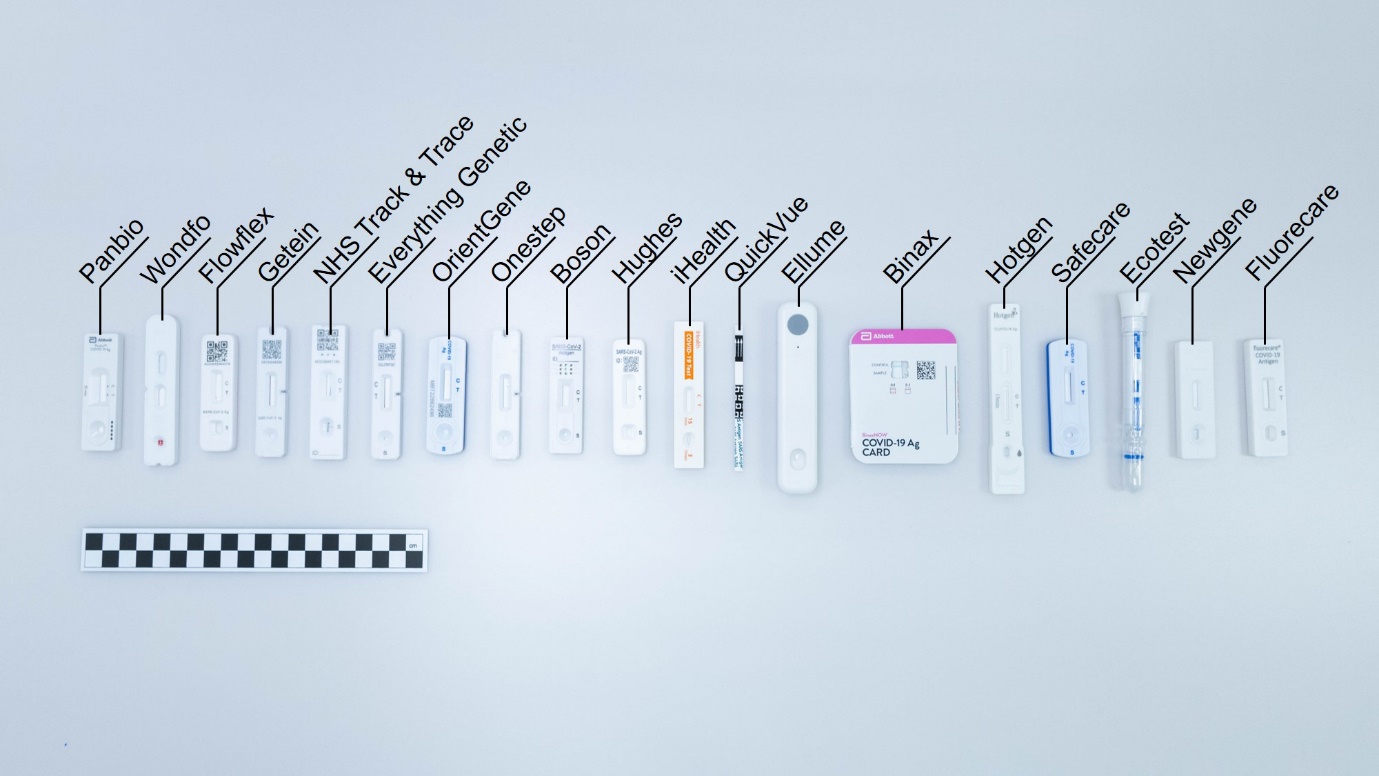

**Figure S.4. Photograph of all swab designs in the collection**

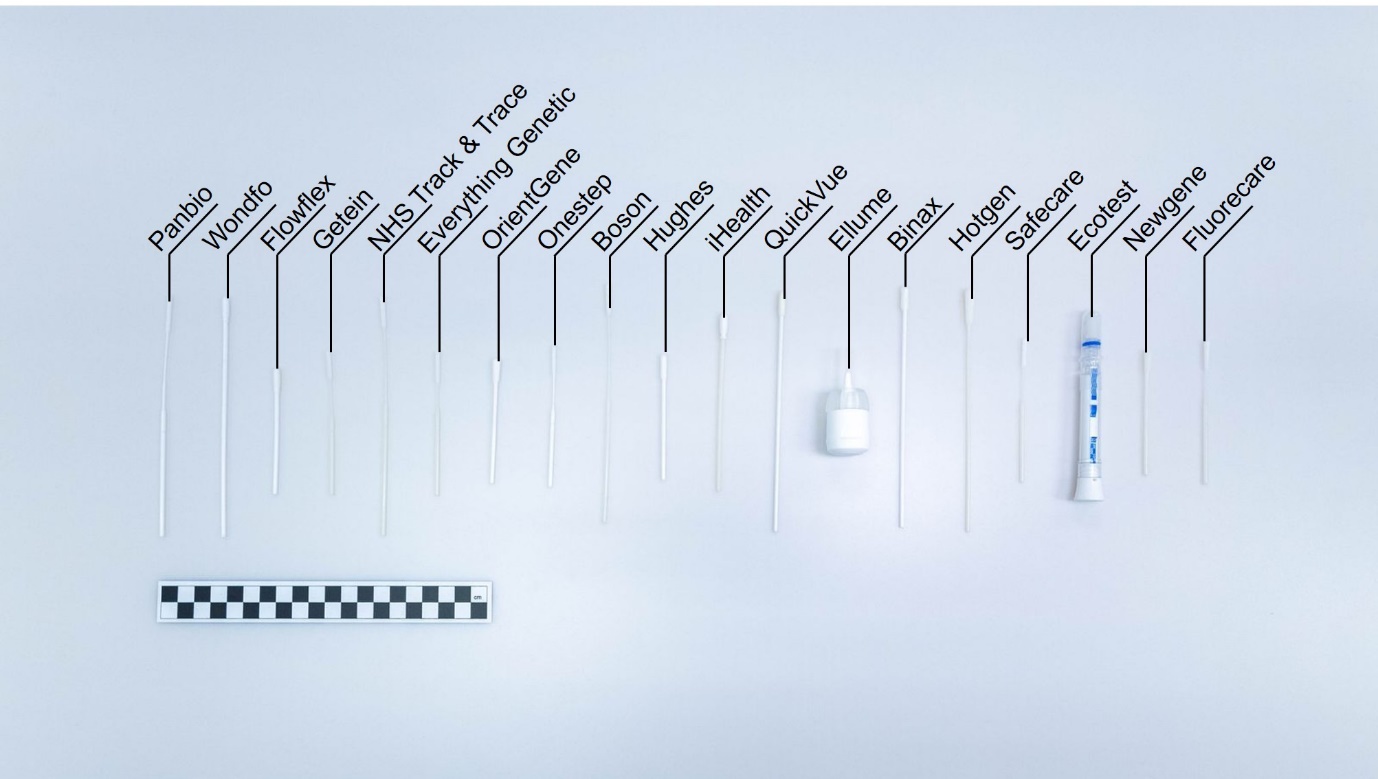

**Figure S.5. Photograph of all reagent tube designs in the collection**
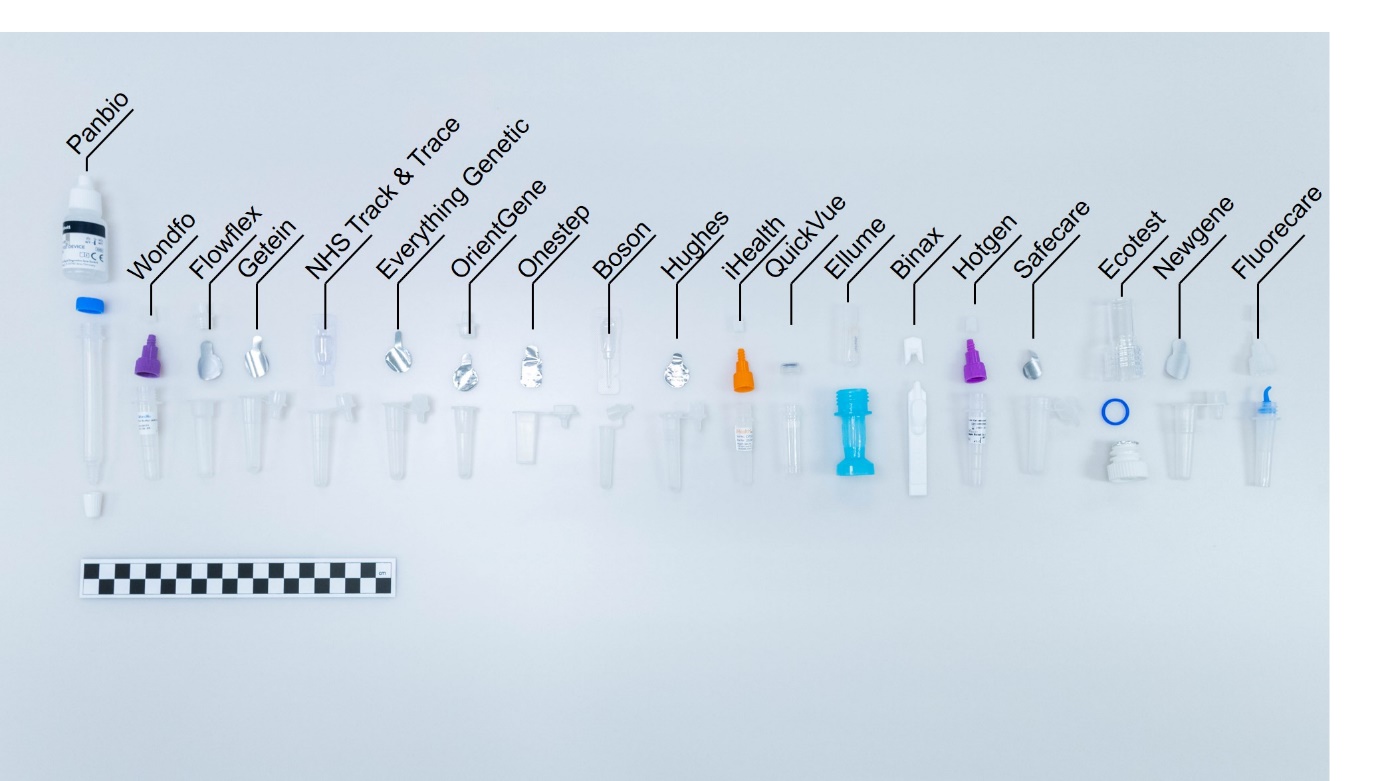
